# Determinants of *de novo* B cell responses to drifted epitopes in post-vaccination SARS-CoV-2 infections

**DOI:** 10.1101/2023.09.12.23295384

**Authors:** Grace E. Quirk, Marta V. Schoenle, Kameron L. Peyton, Jennifer L. Uhrlaub, Branden Lau, Jefferey L. Burgess, Katherine Ellingson, Shawn Beitel, James Romine, Karen Lutrick, Ashley Fowlkes, Amadea Britton, Harmony L. Tyner, Alberto J. Caban-Martinez, Allison Naleway, Manjusha Gaglani, Sarang Yoon, Laura Edwards, Lauren Olsho, Michael Dake, Bonnie J. LaFleur, Janko Ž. Nikolich, Ryan Sprissler, Michael Worobey, Deepta Bhattacharya

## Abstract

Vaccine-induced immunity may impact subsequent *de novo* responses to drifted epitopes in SARS-CoV-2 variants, but this has been difficult to quantify due to the challenges in recruiting unvaccinated control groups whose first exposure to SARS-CoV-2 is a primary infection. Through local, statewide, and national SARS-CoV-2 testing programs, we were able to recruit cohorts of individuals who had recovered from either primary or post-vaccination infections by either the Delta or Omicron BA.1 variants. Regardless of variant, we observed greater Spike-specific and neutralizing antibody responses in post-vaccination infections than in those who were infected without prior vaccination. Through analysis of variant-specific memory B cells as markers of *de novo* responses, we observed that Delta and Omicron BA.1 infections led to a marked shift in immunodominance in which some drifted epitopes elicited minimal responses, even in primary infections. Prior immunity through vaccination had a small negative impact on these *de novo* responses, but this did not correlate with cross-reactive memory B cells, arguing against competitive inhibition of naïve B cells. We conclude that dampened *de novo* B cell responses against drifted epitopes are mostly a function of altered immunodominance hierarchies that are apparent even in primary infections, with a more modest contribution from pre-existing immunity, perhaps due to accelerated antigen clearance.

## Introduction

Within a year of the discovery of SARS-CoV-2 as the etiological agent of COVID-19^1^, highly effective vaccines were developed and administered. Leading this class were the monovalent mRNA vaccines BNT162b2 and mRNA-1273 encoding the ancestral Spike protein, both of which achieved ∼95% efficacies in preventing symptomatic illness^2,3^. Other vaccine platforms also achieved high efficacies, especially against severe illness and hospitalization^4–8^. Since the initial results of these clinical trials, however, the protective capacity of these vaccines has declined^9–12^. This drop in vaccine effectiveness is due to both waning of antibodies and viral evolution and escape from vaccine-induced neutralizing antibodies, which are the best-known correlates of protection^13,14^. While the known genetic diversity of SARS-CoV-2 was quite modest through most of 2020^15^, new variants with enhanced transmissibility and/or neutralizing antibody escape mutations have since emerged and sequentially swept to global dominance^12,16–24^. As of this writing, the dominant circulating variant is Omicron, which comprises sublineages that contain Spike protein mutations located within most known neutralizing antibody epitopes^25^. A key issue that will define both protection against infections and the strategy underlying updates to the vaccines is the extent to which pre-existing vaccine-induced immunity protects against heterologous challenges like Omicron.

B cell responses following mRNA COVID-19 vaccination are characterized by exceptionally long-lived germinal center reactions that persist for months while continuously improving the breadth and affinity of antibodies^26–29^. Cells exiting the germinal center carry affinity-enhancing mutations and can become long-lived antibody-secreting plasma cells or memory B cells^30^. Depending on the subset of memory B cell, re-exposures to antigen trigger differentiation to new plasma cells or germinal center reactions^31–34^. After antigens from infection or vaccine antigens have been cleared, long-lived plasma cells and memory B cells persist to maintain humoral immunity.

While these features protect against homologous SARS-CoV-2 infections, it is more difficult to predict the nature of responses to subsequent heterologous infections or vaccines. Due to their expanded pre-existing numbers and intrinsic signaling and transcriptional differences relative to naive B cells, memory B cells rapidly mount responses that are of greater magnitude than those of naïve primary responses^35–40^ to either initial infection or vaccination. Because of these properties, memory B cells that react to epitopes conserved between the original and secondary challenges could dominate the response to heterologous infections or vaccines^41–43^. If antigen and T cell help are limiting, memory B cells might then outcompete naive B cells and new primary antibody responses aimed at the new variant-specific epitopes. This phenomenon, known as antigenic imprinting or “original antigenic sin”^44^, can be beneficial if antibodies against the conserved epitopes are protective. However, recall responses to heterologous pathogens can potentially be neutral or even detrimental if antibodies targeting these conserved epitopes are not protective and variant-specific primary responses are competitively inhibited^45^. As an example of the phenomenon, pre-existing common coronavirus-specific memory B cells compose a majority of the early response to SARS-CoV-2, but primary responses to epitopes unique to SARS-CoV-2 are readily observed later^26,46,47^. Whether common coronavirus immunity is helpful, harmful, or neutral for *de novo* responses to SARS-CoV-2 is unknown.

In influenza infections, antigenic imprinting has been proposed to explain the age-associated differential in morbidity and mortality based on influenza subtype exposure history^48–51^. The various hemagglutinin (HA) subtypes of influenza A virus fall into one or the other of two phylogenetically distinct HA “groups” (group 1 or group 2). Individuals have the highest antibody titers against influenza strains encountered early in life, and they experience enhanced protection against influenza strains that are within the same HA group as their primary infection strain compared to heterosubtypic infections from the group that is mismatched to their first childhood infection. Previous work has shown that childhood exposure to H1N1 (group 1 hemagglutinin (HA)) affords protection against other group 1 HAs, such as H5N1. The same is true for individuals with group 2 HAs, whereby childhood H3N2 infection affords protection against H7N9. Conversely, individuals with group 1 imprinting experience an increase in mortality when faced with a group 2 influenza virus infection, such as that observed for H7N9 infections^48,51^.

Though pre-existing immunity can certainly impact primary responses to heterologous antigens, other mechanisms can also limit antibody responses to drifted epitopes. Epitopes that were previously immunodominant for antibody responses do not necessarily remain so once mutated, irrespective of prior immunity^52^. There are several possible mechanistic reasons why not all epitopes are equal for antibody responses. Factors such as naïve antigen-specific B cell precursor frequency and avidity vary greatly across epitopes, which in turn correlate with their relative contribution to the subsequent response^53–57^. Some epitopes can also be biophysically challenging for antibody binding, such as those sterically blocked by glycan shields or appearing as non-complex ‘smooth’ surfaces to B cells^58,59^. Further, epitopes that mimic self-antigens also elicit poor responses due to tolerance mechanisms that remove or hamper B cells from the repertoire that could otherwise respond^60–63^. Finally, V gene usage during V(D)J recombination that gives rise to B cell receptors is uneven, as some segments are more heavily utilized than others^64,65^. In turn, this can create ‘holes’ in the repertoire, rendering some epitopes poorly immunogenic^66^. As SARS-CoV-2 variants of concern accumulate mutations in antigenic regions, immunodominance might change in ways that limit responses to drifted epitopes, with or without prior immunity. Thus, it has remained difficult to examine the degree to which infrequent *de novo* variant-specific responses in post-vaccination infections and heterologous boosters are due to changes in immunodominance, antigenic imprinting, or some combination of both^67–75^.

Antigenic imprinting has remained nearly impossible to quantify directly and instead has predominantly relied on historical epidemiological data to make inferences about biological mechanisms that produce the documented patterns^48,76,77^. The COVID-19 pandemic presents a unique opportunity to address these questions: it has encompassed adults with known infection histories and monovalent vaccines that induce well characterized B cell responses^78–81^ and the emergence of antigenically distinct viral variants^25,82^. Yet, as immunological histories become more complex and with very few immunologically naïve adults remaining^83,84^, the Omicron BA.1 (BA.1, for short) wave likely represented the final opportunity to recruit robust cohorts of individuals that meet the key experimental and control criteria. Through voluntary saline-gargle PCR testing of University of Arizona students, staff, and faculty as part of COVID mitigation efforts on campus from August 2020 to July 2023; serological testing at 17 University of Arizona-managed sites across the state of Arizona; and two CDC-funded cohorts of essential workers, Arizona Healthcare, Emergency Response, and Other Essential Workers Surveillance (AZ HEROES)^85^ and Research on Epidemiology of SARS-CoV-2 in Essential Response Personnel (RECOVER)^86^, we recruited unvaccinated individuals who had recovered from primary Delta (B.1.617.2 or B.1.617.2-like) or BA.1 (B.1.1.529 or B.1.1.529-like) infections. These cohorts allowed us to characterize the immunodominance hierarchies for both Delta and BA.1 variants and directly compare the specificity of antibody responses in unvaccinated individuals infected by the antigenically drifted viral variants to those generated by post-vaccination infection by Delta or BA.1. In doing so, we were able to quantify the impact of antigenic imprinting on *de novo* responses to drifted epitopes.

## Results

From our voluntary on-campus testing program at the University of Arizona, we recruited 37 participants who had tested positive for SARS-CoV-2 infections between July 1, 2021 and December 1, 2021 despite completion of the primary vaccine series of monovalent BNT162b2 or mRNA-1273 prior to infection (described in detail in Methods section). We also recruited 12 individuals who tested positive during this period but had not received any COVID-19 vaccines. Symptoms reported by participants following infections were similar between primary and post-vaccination infections; none required hospitalization. A slightly larger portion of post-vaccination infections were asymptomatic relative to primary infections (**Figure S1A**), and in general, the duration of symptoms was significantly shorter in this group relative to those who were unvaccinated at the time of infection (**Figure S1B**). All recruited individuals who tested positive by RT-qPCR and had sufficient sequence coverage to assign a lineage had sequences confirmed to be Delta (**Figure S2A**). During this period, the Delta variant represented 100% of PCR+ samples on campus that could be assigned a PANGO-lineage^87^, as determined through viral sequencing of all remnant samples below a Ct value of 35 (**Figure S2B)**. We also selected 71 serum samples as part of our statewide antibody testing initiative^88^ from vaccinated participants who had no self-reported prior SARS-CoV-2 infections. This cohort was chosen based on matching for age, sex, and time post-vaccination with our post-vaccination infection group. Characteristics of the cohorts are listed in **Table 1**.

**Table 1.** Characteristics of cohorts Interquartile range (IQR) is listed in parentheses unless otherwise stated in the table.

|  | vaccinated only<br>(statewide antibody<br>testing initiative)<br>(n=74) | primary Delta<br>(n=12) | post-vaccination<br>Delta (n=37) | vaccinated only<br>(TATS) (n=62) | primary<br>BA.1 (n=69) | post-vaccination<br>BA.1 (n=62) |
| --- | --- | --- | --- | --- | --- | --- |
| **Age**<br>Mean (s.d.) | 38.0 (32.0, 54.0) | 21.9 (20.2, 40.7) | 23.3 (18.6, 65.8) | 31.9 (18.6, 65.0) | 44 (25, 62) | 40 (19, 71.5) |
| **Sex** |  |  |  |  |  |  |
| Male | 22 (32%) | 5 (42%) | 8 (22%) | 24 (39%) |  | 24 (39%) |
| Female | 52 (68%) | 7 (58%) | 29 (78%) | 37 (60%) |  | 34 (55%) |
| **Prior COVID infection** |  |  |  |  |  |  |
| Yes |  | 12 (100%) | 37 (100%) |  | 69 (100%) | 64 (100%) |
| No | 74 (100%) |  |  |  |  |  |
| Time since COVID infection |  | 67.5 days (32, 99.3) | 71.5 days (48.5, 89.5) |  | 40 days (31, 44.5) | 54.4 days (34.5, 71) |
| paired pre- and post- infection samples |  |  |  |  | 10 | 21 |
| Time from vaccination to pre- infection draw |  |  |  |  |  | 138 (32.5, 217) |
| Time from pre- infection draw to infection |  |  |  |  | 73.3 days (30, 99) | 112 days (33, 187) |
| Time from infection to post- infection draw |  |  |  |  | 42.7 days (29.5, 47.5) | 44.8 days (34, 49.3) |
| **COVID Vaccination # of shots** | 37 | 0 | 37 | 62 | 0 | 62 |
| 2 | 37 (100%) |  | 37 (100%) | 16 (26%) |  | 13 (20%) |
| 3 |  |  |  | 46 (74%) |  | 15 (23%) |
| 4 |  |  |  |  |  | 3 (5%) |
| time since vaccination | 135.6 days (126, 270) |  | 273.9 days (56, 317) | 176.0 days (113.5, 188) |  | 192.2 days (107, 302.3) |

Participants provided blood samples at an average of 75 days (IQR for primary and post-vaccination infections = 45.8 days, 97.3 days; **Table 1**) after testing positive for SARS-CoV-2 infections and at an average of nine months (IQR for vaccinated only and post-vaccination infections = 56 days, 317 days; **Table 1**) after their last vaccine dose. Using plasma from these samples, we first performed live virus neutralization assays on both an early-pandemic virus representative, (WA-1, from January 2020) or on the antigenically drifted Delta variant. Against both WA-1 and Delta, post-vaccination Delta infections led to significantly higher titers of neutralizing antibodies than both primary infections and vaccinated only controls (**Figure 1A**), indicating a robust recall response.

**Figure 1.**
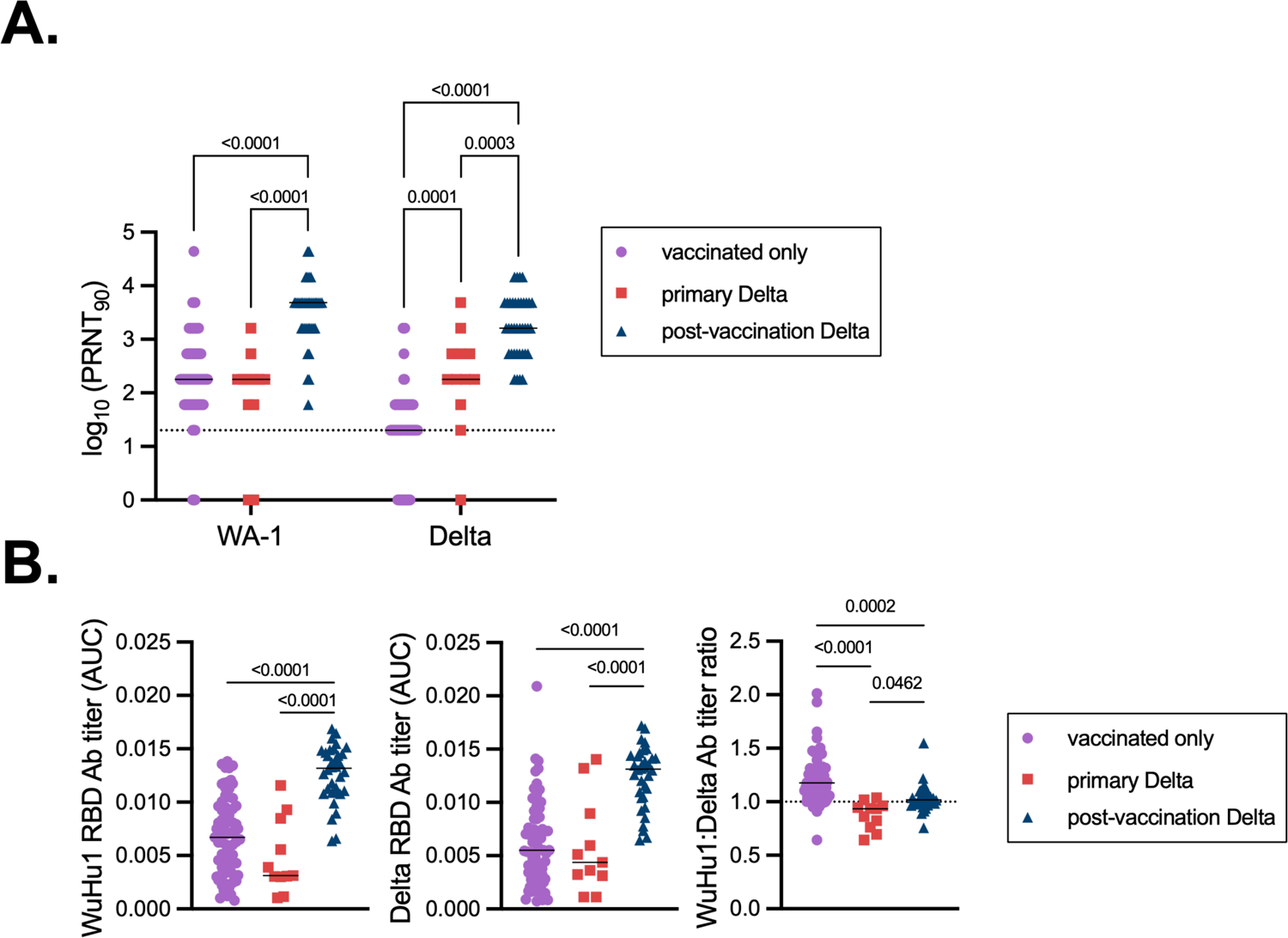
Primary and recall antibody responses to Wuhan and Delta strains of SARS-COV-2. **(A)** Virus neutralization assays were performed using the WA-1 and Delta isolates of SARS-CoV-2. Serial 1:3 dilutions of serums were performed and tested for the ability to prevent plaque formation on Vero cells. The lowest concentration capable of preventing more than 90% of plaques was considered to be the PRNT_90_ value. Each symbol represents an individual. Two-sided P values from t-test statistics were calculated for pairwise differences using two-way ANOVA. Post hoc testing for multiple comparisons between draws was performed using Tukey’s multiple comparisons test. P values greater than 0.05 are not depicted. (**B)** Quantitative titers of WuHu1- and Delta RBD-specific antibodies. Serum was initially diluted 1:60, serially diluted 1:3, assessed by ELISA for binding to the listed antigens, and area under the curve (AUC) values were calculated. Each symbol represents an individual. WuHu1 AUC values were divided by their Delta AUC titer in the same individual to calculate a WuHu1:Delta RBD ratio in the rightmost panel. Two-sided P values from t-test statistics were calculated for pairwise differences using one-way ANOVA. Post hoc testing for multiple comparisons between draws was performed using Tukey’s multiple comparisons test. P values greater than 0.05 are not depicted.

Elevated neutralizing antibody titers in post-vaccination Delta infections could arise from both memory B cell responses to conserved neutralizing epitopes and primary responses against new variant-specific epitopes. To begin to determine the relative specificities of antibodies following Delta infections, we performed ELISAs to measure the magnitude of the antibody response against Wuhan/Hu1/2019 (hereafter WuHu1) and Delta Spike antigens. WuHu1 was sampled in December 2019 and is the SARS-CoV-2 reference sequence; its Spike amino acid sequence is identical to that of WA-1. We first measured antibodies that bound the receptor binding domain (RBD), as most neutralizing antibodies target this region^89,90^. Post-vaccination Delta infections led to elevated RBD-binding antibody titers, both against WuHu1 and Delta, relative to vaccination only and primary Delta infection controls (**Figure 1B**), again confirming a robust recall response. As expected, vaccination-only controls showed slightly elevated titers of WuHu1 RBD-binding antibodies relative to Delta RBD antibodies (**Figure 1B, right panel**). Reciprocally, primary Delta infections led to a skewing towards Delta RBD-binding antibodies (**Figure 1B, right panel**). Post-vaccination-Delta infections led to an even ratio of WuHu1:Delta RBD-binding antibodies (**Figure 1B, right panel**), similar to prior studies^91^. Aside from the RBD, neutralizing antibodies can also bind other regions of the S1 domain of Spike^92–94^. As with RBD, post-vaccination Delta infections led to an even ratio of antibodies that bound WuHu1 and Delta S1 relative to vaccination alone or primary Delta infections (**Figure S3**).

To more directly assess antibody specificities with single cell resolution in post-vaccination Delta infections, memory B cells using WuHu1 S1 and Delta S1 antigen tetramers were quantified by flow cytometry. We focused our analysis on the isotype-switched CD27+ subset (**Figure 2A and Figure S4**), since few Spike-specific cells are observed in other memory B cell subsets^95^. Memory B cells that bound Delta S1 only were observed in both primary infections and in post-vaccination Delta infections, suggesting that in both cases, *de novo* responses aimed at variant-unique epitopes were mounted (**Figure 2A-B**). However, the proportions of these cells in PBMCs were slightly reduced in post-vaccination Delta infections relative to primary Delta infections (**Figure 2B**). Reciprocally, cross-reactive memory B cells that bound both WuHu1 S1 and Delta S1 were elevated in post-vaccination Delta infections relative to primary Delta infections (**Figure 2B**), consistent with a robust recall response and antigenic imprinting, though for a subset of individuals this appears to be more modest. In both primary and post-vaccination Delta infections, memory B cells that bound Delta S1 uniquely were rare relative to cross-reactive cells that bound both WuHu1 and Delta S1 (**Figures 2A-B**). Although these data suggest that pre-existing immunity limits new primary responses, cross-reactive and Delta-specific memory B cells were positively correlated in post-vaccination Delta infections (**Figure 2C**), arguing against a mechanism of competitive inhibition between these two cellular compartments.

**Figure 2.**
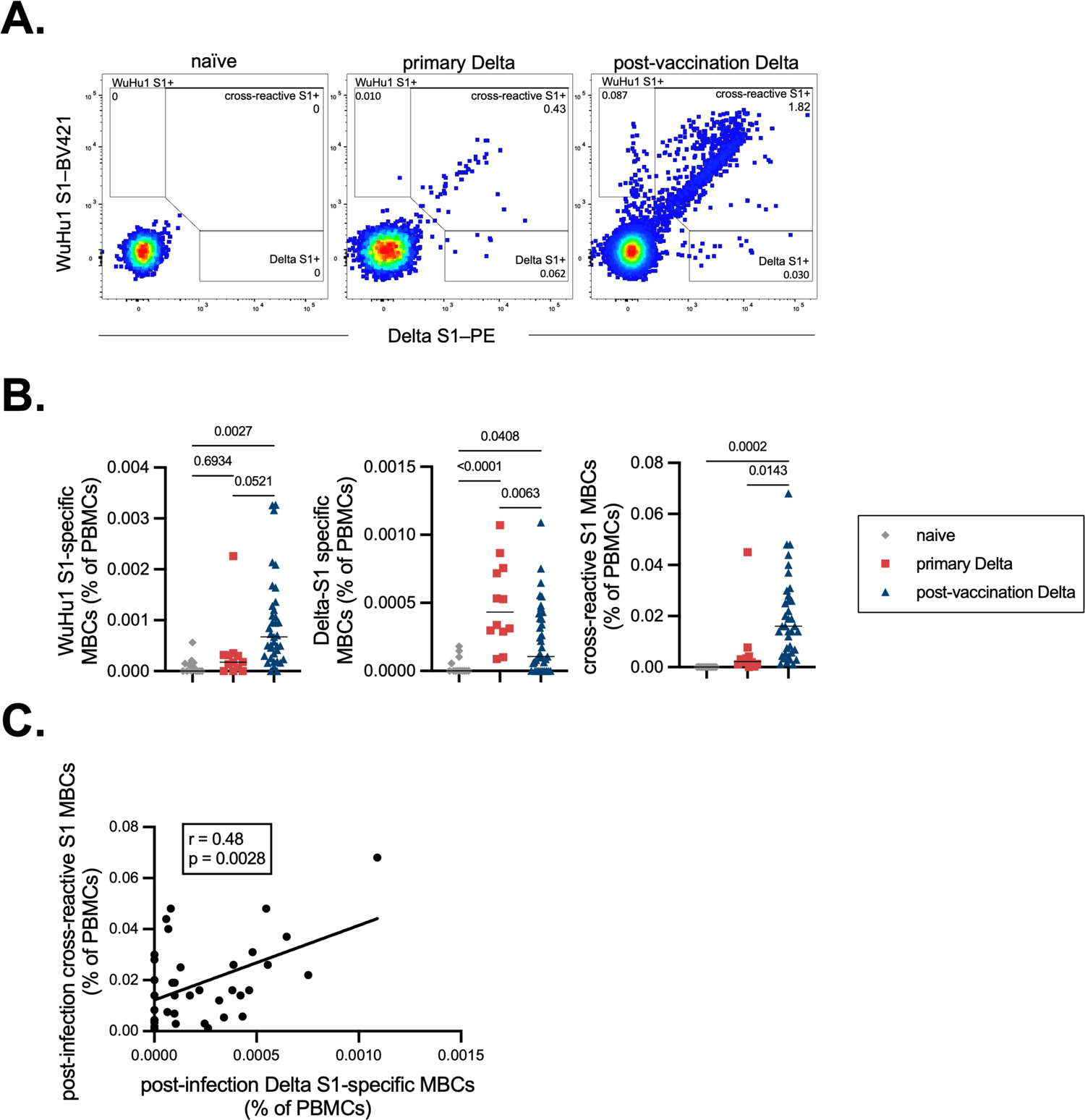
WuHu1 and Delta Memory B cell flow cytometric analysis and quantification. **(A)** Representative flow cytometric plots of Wuhu1 and Delta S1-specific memory B cells (full gating strategy shown in Figure S2) in naïve, primary Delta infection, and post-vaccination Delta infection cohorts. Cells that bind both WuHu1 S1 and Delta S1 are annotated as cross-reactive S1+, whereas cells that bind only WuHu1 S1 or Delta S1 are annotated as WuHu1 S1+ or Delta S1+, respectively. **(B)** Quantification of isotype-switched memory B cells as a percentage of total PBMCs for Wuhu1 S1+, Delta S1+ and cross-reactive S1+ specificities for each cohort of SARS-CoV-2 immune histories. Each symbol represents an individual. Two-sided P values from t-test statistics were calculated for pairwise differences using one-way ANOVA. Post hoc testing for multiple comparisons between draws was performed using Tukey’s multiple comparisons test. P values greater than 0.05 are not depicted. **(C)** Correlation of post-infection cross-reactive S1 MBCs (calculated as in Figure 2B) plotted against the frequency of post-infection Delta S1-specific MBCs (calculated as in Figure 2B) in individuals that experienced a post-vaccination Delta infection. Pearson correlation analysis was performed.

The RBD of Delta contains two non-synonymous point mutations that deviate from the vaccine sequence: T478K and L452R. The L452R mutation in particular leads to neutralizing antibody escape^90,96–98^. To estimate the epitope preferences of serum antibodies further, we produced a Delta RBD protein in which R452 was reverted to L452. Vaccination led to a response that was skewed toward the L452-containing RBD (**Figure 3A**, compare to **Figure 1B, middle panel**), confirming the strong antibody bias and immunodominance of this epitope reported previously^99^. Yet reciprocal skewing to R452-containing RBD was not observed in primary Delta infections, suggesting that a new immunogenic epitope is not created by this mutation (**Figure 3A**). Post-vaccination Delta infections led to a relatively even ratio of antibodies that bound Delta-L452 to those that reacted to Delta-R452 **(Figure 3A**), perhaps due to boosted levels of antibodies that bound other conserved sites on RBD and the T478K epitope. We also produced chimeric WuHu1 S1 proteins in which the Delta N-terminal domain (NTD) supersite mutations (T19R, G142D, E156-, F157-, R158G) were introduced onto a WuHu1 background^92–94^. Vaccination only controls showed a relatively even distribution of antibodies that bound WuHu1 S1 and Delta NTD-WuHu1 S1 (**Figure 3B** compare to **Figure S3, left panel**). Primary Delta infections, however, were subtly but significantly skewed towards the Delta NTD (**Figure 3B**). Together, these data demonstrate a shifting of immunodominance profiles, even in the absence of prior SARS-CoV-2 immunity.

**Figure 3.**
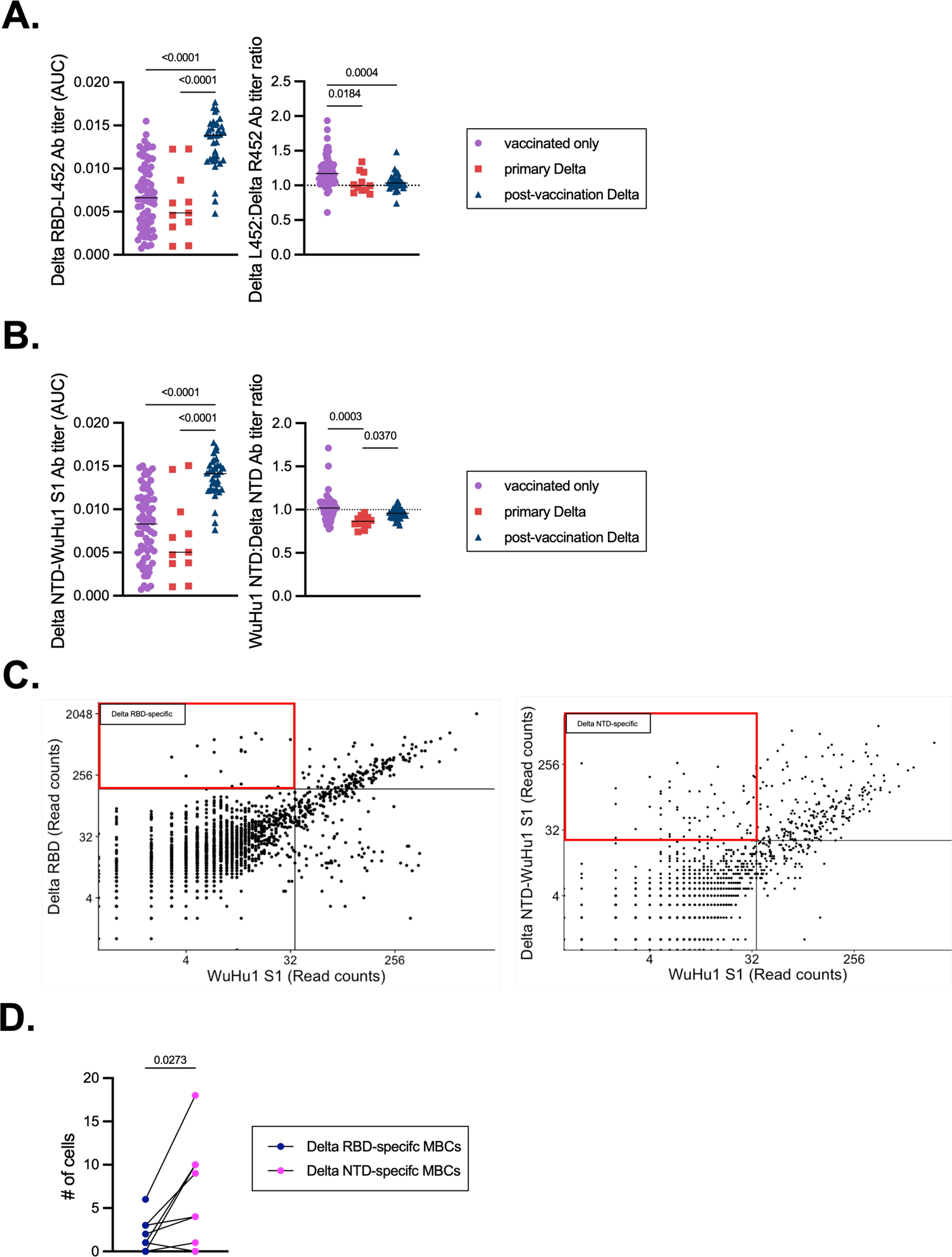
Epitope-specific quantification of Delta RBD- and Delta NTD-specific antibodies and memory B cells. (**A)** A chimeric protein (Delta RBD-L452) was generated in which R452 was reverted to the ancestral L452. ELISAs were used to quantify serum antibodies that bound to Delta RBD-L452 in each cohort. Delta RBD-L452 AUC titers were divided by Delta RBD titers (Figure 1B) in the same individuals to calculate a L452:R452 titer ratio. Each symbol represents an individual. Two-sided P values from t-test statistics were calculated for pairwise differences using one-way ANOVA. Post hoc testing for multiple comparisons between draws was performed using Tukey’s multiple comparisons test. P values greater than 0.05 are not depicted. **(B)** A chimeric protein (Delta NTD-WuHu1 S1) was generated in which Delta NTD mutated epitopes (T19R, G142D, E156-, F157-, R158G) were incorporated into the otherwise WuHu1 S1 backbone. ELISAs were used to quantify serum antibodies that bound to Delta NTD-WuHu1 S1 in each cohort. Delta RBD-L452 AUC titers were divided by their Delta RBD (Supplemental Fig 1A) titer to calculate a WuHu1 NTD:Delta NTD titer ratio. Each symbol represents an individual. Two-sided P values from t-test statistics were calculated for pairwise differences using one-way ANOVA. Post hoc testing for multiple comparisons between draws was performed using Tukey’s multiple comparisons test. P values greater than 0.05 are not depicted. **(C)** LIBRA-seq plots of isotype-switched memory B cells enriched for Spike-binding specificities from primary Delta infections. Read count thresholds to determine positivity were set using samples in which cells lacking Spike-binding specificities were sorted and sequenced. Plots are concatenated from ten individuals. **(D)** Quantification of Delta RBD-specific and Delta NTD-specific memory B cells (MBCs) in individuals that experienced a primary Delta infection. Lines connect specificities within the same individual. Delta RBD-specific cells were classified by cells that had Delta RBD read counts of greater than 160 and WuHu1 S1 read counts of less than 35. Delta NTD-specific cells were classified by cells that had Delta NTD-WuHu1 S1 read counts of greater than 23 and WuHu1 S1 read counts of less than 35. Two-sided P values were calculated for pairwise differences using paired t-tests.

To more precisely measure clonal shifts in antibody specificities and immunodominance than can be achieved by serological assays, we performed LIBRA-seq using PBMC samples from primary and post-vaccination Delta infections^100^. Streptavidin-phycoerythrin (PE) tetramers were constructed using WuHu1 S1, Delta S1, Delta RBD, Delta RBD-L452, and Delta NTD-WuHu1-S1, as described in **Figures 1, 3** and **S3**, each carrying unique oligonucleotide barcodes. PE-binding memory cells were then enriched and subjected to scRNA/V(D)J-seq. Consistent with our serological data (**Figure 3A-B**), we observed few memory B cells that bound Delta RBD- and NTD-specific epitopes (**Figure 3C**) in primary Delta infections and post-vaccination Delta infections (**Figure S5A**). A clear preference for Delta-unique epitopes in the NTD relative to the RBD was observed within individuals that had experienced a primary Delta infection (**Figure 3D**). Within each group, we did not observe any clear differences in epitope-dependence of somatic mutation frequencies in memory B cells (**Figure S5B**). We did, however, observe a greater frequency of somatic mutations in Spike-specific memory B cells in the post-vaccination Delta infection cohort relative to primary Delta infections (**Figure S5C**). Together, these data suggest a marked shift in antibody specificities in primary Delta variant infections relative to WuHu1 Spike. This explains in part why responses to at least some drifted epitopes are not observed, irrespective of prior vaccination.

During the course of this work, the heavily mutated Omicron (BA.1) variant rapidly overtook Delta and swept to global dominance. To define post-vaccination BA.1 responses, we recruited individuals from our voluntary on-campus testing program who had tested positive for SARS-CoV-2 between January 1 and March 31, 2022, with the expectation that primary responses would be robust against this more antigenically distant variant^101^. All individuals for this study who tested positive by PCR had sequences confirmed to be BA.1 (**Figure S2A**). Individuals with a SARS-CoV-2 infection caused by a Delta variant or other Omicron sublineages were excluded from the study. Viral genome sequencing of all remnant PCR+ samples on campus during this period below a Ct value of 35 demonstrated that 93.7% of samples that could be assigned a PANGO-lineage^87^ were caused by the BA.1 sublineage of Omicron (**Figure S2B**). To obtain controls for this cohort, some of whom had received 3 doses of mRNA vaccines, we also recruited a new group of vaccinated individuals who had never tested positive in our voluntary university testing system and reported no known prior SARS-CoV-2 infections. After testing plasma for nucleocapsid antibodies as a marker of prior infection, samples from 5 individuals with titers well above the mean values seen in verified infections were excluded from further consideration (**Figure S6**). Relative to both primary and post-vaccination Delta infections, post-vaccination BA.1 infections generally led to fewer symptoms such as wet cough (**Figure S1A**) and shorter duration of symptoms (**Figure S1B**).

We were unable to recruit any unvaccinated individuals on campus who had experienced BA.1 infections. However, we were able to obtain serum and, for a subset, PBMC samples from a separate study from the Centers for Disease Control and Prevention HEROES and RECOVER projects^85^, in which 53 individuals met these criteria (**Table 1**). Neutralizing antibody titers were skewed towards WA-1 in individuals who had been vaccinated but not infected (**Figure 4A**). Post-vaccination BA.1 infections led to significantly higher neutralizing antibody titers against BA.1 compared to both vaccinated controls who had not been infected and primary infections (**Figure 4A**), consistent with a memory B cell recall response.

**Figure 4.**
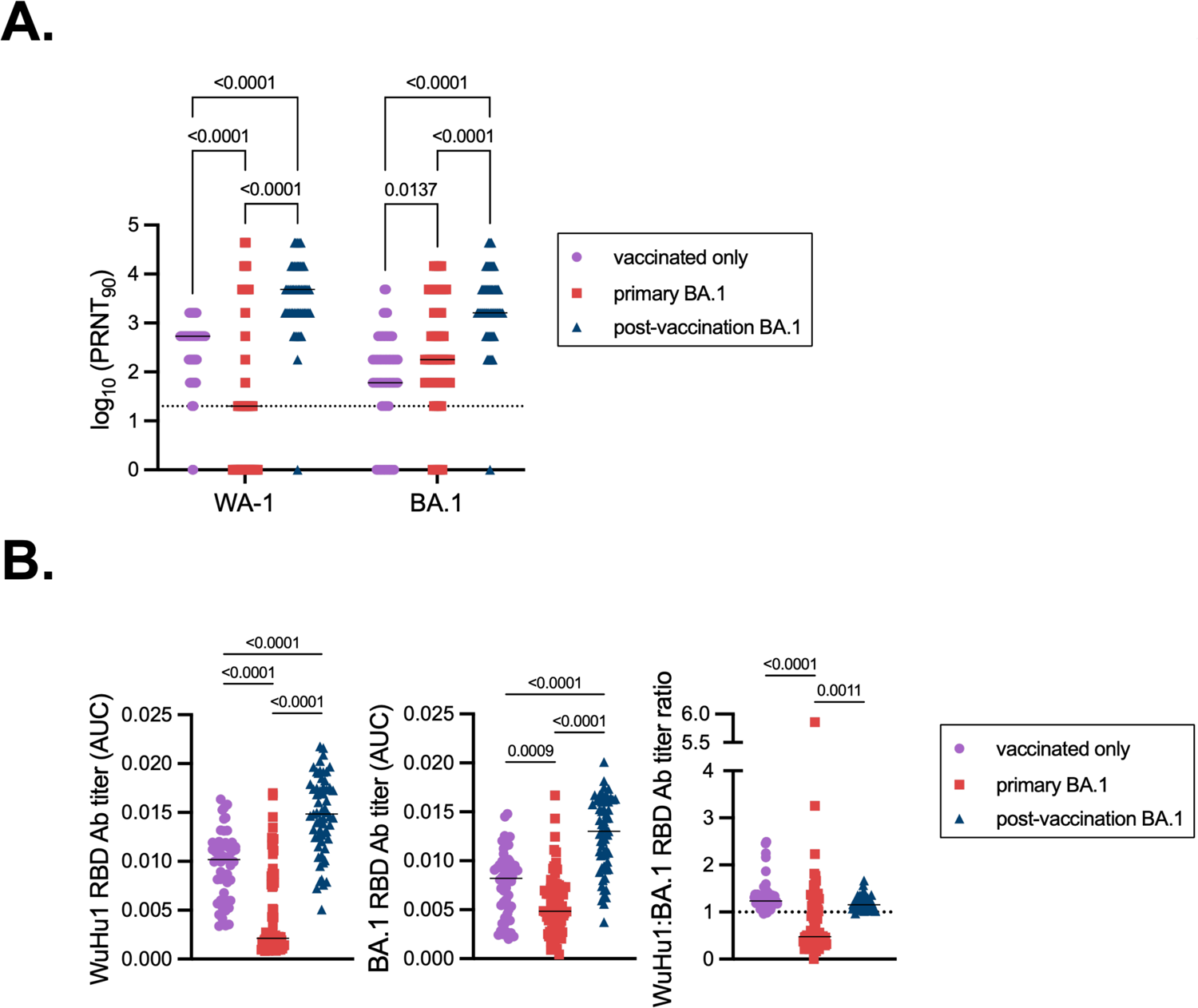
Primary and recall antibody responses to Wuhan and BA.1 strains of SARS-COV-2. **(A)** Virus neutralization assays were performed using the WA-1 and BA.1 isolates of SARS-CoV-2. Serial 1:3 dilutions of serums were performed and tested for the ability to prevent plaque formation on Vero cells. The lowest concentration capable of preventing more than 90% of plaques was considered to the PRNT_90_ value. Each symbol represents an individual. Two-sided P values from t-test statistics were calculated for pairwise differences using two-way ANOVA. Post hoc testing for multiple comparisons between draws was performed using Tukey’s multiple comparisons test. P values greater than 0.05 are not depicted. **(B)** Quantitative titers of Wuhu1 and BA.1 RBD antibodies. Serum was initially diluted 1:60, serially diluted 1:3, assessed by ELISA for binding to the listed antigens, and area under the curve (AUC) values were calculated. Each symbol represents an individual. WuHu1 AUC values were divided by their BA.1 RBD AUC titer in the same individual to calculate a ratio in the rightmost panel. Two-sided P values from t-test statistics were calculated for pairwise differences using one-way ANOVA. Post hoc testing for multiple comparisons between draws was performed using Tukey’s multiple comparisons test. P values greater than 0.05 are not depicted.

We next examined binding antibody titers against WuHu1 or BA.1 RBD. Post-vaccination BA.1 infections led to increased levels of RBD-binding antibodies, both for WuHu1 and BA.1, relative to the vaccinated only control cohort and primary BA.1 infections (**Figure 4B, left and middle panels**). Vaccination alone led to greater RBD titers against BA.1 than did primary BA.1 infections, despite the many mismatches in sequence (**Figure 4B, middle panel**). As expected, antibodies from vaccinated only individuals were skewed towards WuHu1 relative to BA.1 RBD (**Figure 4B, right panel**). Of the few antibodies induced by primary BA.1 infections, we observed a skewing of specificities towards BA.1 RBD (**Figure 4B, right panel**). Ratios of WuHu1 and BA.1 RBD-binding antibodies in post-vaccination BA.1 infections more closely resembled vaccinated controls than primary BA.1 infections (**Figure 4B, right panel**).

To further evaluate the specificities of antibody responses in post-vaccination BA.1 infections, we again used antigen tetramers to identify RBD-specific memory B cells (**Figures S7 and 5A**). As expected, primary BA.1 infections generated a lower frequency of WuHu1 RBD-specific memory B cells compared to vaccinated controls (**Figure 5B, left panel**). Unexpectedly, BA.1-specific RBD memory B cells were not consistently detectable above background in any experimental group, even primary BA.1 infections (**Figure 5B, middle panel**). These data seem to differ from the modest skewing of the serological response seen above in primary BA.1 infections (**Figure 4B, right panel**), but can potentially be explained by low overall responses and prior studies that observed only partial overlap between memory B and antibody-secreting plasma cell specificities and repertoires^43,102,103^. Instead, most RBD-specific memory B cells from all cohorts were cross-reactive against WuHu1 and BA.1 RBD (**Figure 5A, 5B, right panel**). Primary BA.1 infections produced numerically fewer cross-reactive RBD memory B cells than did post-vaccination BA.1 infections (**Figure 5B, right panel**).

**Figure 5.**
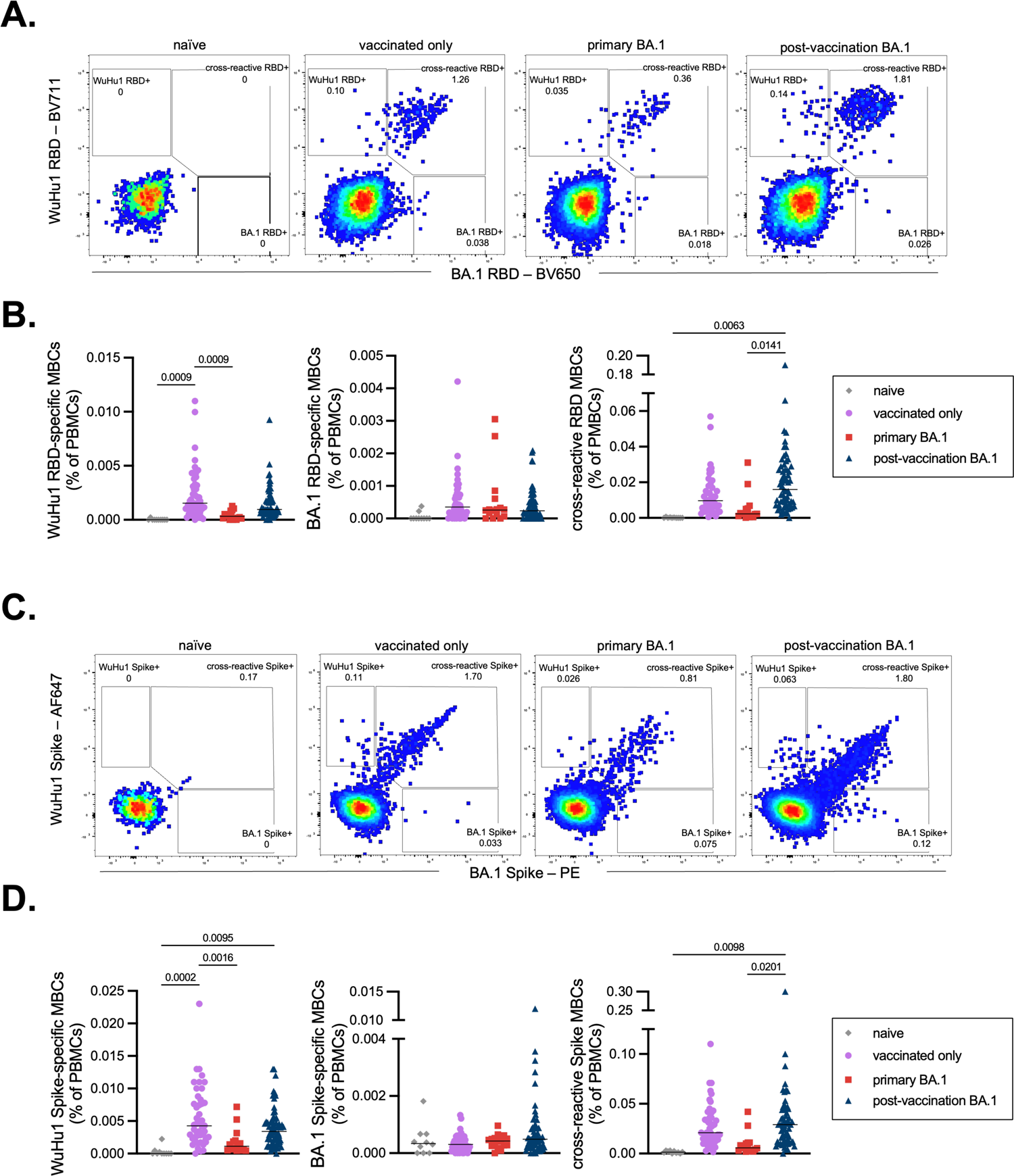
WuHu1 and BA.1 Memory B cell flow cytometric analysis and quantification. **(A)** Representative flow cytometric plots of Wuhu1 and BA.1 RBD-specific memory B cells (full gating strategy shown in Figure S3) in naïve, vaccinated only, primary BA.1 infection, and post-vaccination BA.1 infection cohorts. Cells that bind both WuHu1 RBD and BA.1 RBD are annotated as cross-reactive RBD+, whereas cells that bind only WuHu1 RBD or BA.1 RBD are annotated as WuHu1 RBD+ or BA.1 RBD+, respectively. **(B)** Quantification of isotype-switched memory B cells for Wuhu1 RBD+, BA.1 RBD+ and cross-reactive RBD+ specificities for each cohort of SARS-CoV-2 immune histories. Each symbol represents an individual. Two-sided P values from t-test statistics were calculated for pairwise differences using one-way ANOVA. Post hoc testing for multiple comparisons between draws was performed using Tukey’s multiple comparisons test. P values greater than 0.05 are not depicted. **(C)** Representative flow cytometric plots of Wuhu1 and BA.1 Spike-specific memory B cells (full gating strategy shown in Figure S3) in naïve, vaccinated only, primary BA.1 infection, and post-vaccination BA.1 infection cohorts. Cells that bind both WuHu1 RBD and BA.1 Spike are annotated as cross-reactive Spike+, whereas cells that bind only WuHu1 Spike or BA.1 Spike are annotated as WuHu1 Spike+ or BA.1 Spike+, respectively. **(D)** Quantification of isotype-switched memory B cells for Wuhu1 Spike+, BA.1 Spike+ and cross-reactive Spike+ specificities for each cohort of SARS-CoV-2 immune histories. Each symbol represents an individual. Two-sided P values from t-test statistics were calculated for pairwise differences using one-way ANOVA. Post hoc testing for multiple comparisons between draws was performed using Tukey’s multiple comparisons test. P values greater than 0.05 are not depicted.

Given that the overall antibody and memory B cell response to BA.1 RBD was quite modest (**Figures 4B, 5A-B**), we employed tetramers of full-length Spike trimers of WuHu1 and BA.1 Spike to capture a greater breadth of memory B cell specificities than could be observed with RBD tetramers (**Figures 5C**). WuHu1-specific memory B cells were observed in vaccinated controls and post-vaccination BA.1 infections, but not after primary BA.1 infections (**Figures 5D, left panel**). We again failed to consistently observe BA.1-specific memory B cells in any of the groups, including primary BA.1 infections, though a subset of post-vaccination BA.1 infections did appear to generate such cells well above background levels (**Figure 5D, middle panel**). As with RBD, cross-reactive Spike-specific memory cells were significantly elevated in post-vaccination BA.1 infections relative to primary BA.1 infections, but not relative to vaccinated only controls (**Figure 5D, right panel**). Cross-reactive memory B cells composed by far the largest portion of SARS-CoV-2 specific responses within all experimental groups (**Figure S8**).

For a subset of primary BA.1 and post-vaccination BA.1 cohorts, we obtained samples which enabled us to quantify WuHu1, BA.1, and cross-reactive Spike- and RBD-specific memory B cell frequencies before and after BA.1 infection. Irrespective of vaccination status, memory B cells that were either WuHu1- or BA.1-RBD-specific increased in frequency for only a subset of individuals after BA.1 infection (**Figure 6A, left and middle panels**). However, cross-reactive RBD memory B cells consistently and significantly increased after both primary and post-vaccination BA.1 infections (**Figure 6A, right panel**). The frequency of cross-reactive Spike memory B cells also significantly increased after primary BA.1 infections (**Figures 6B, right panel**).

**Figure 6.**
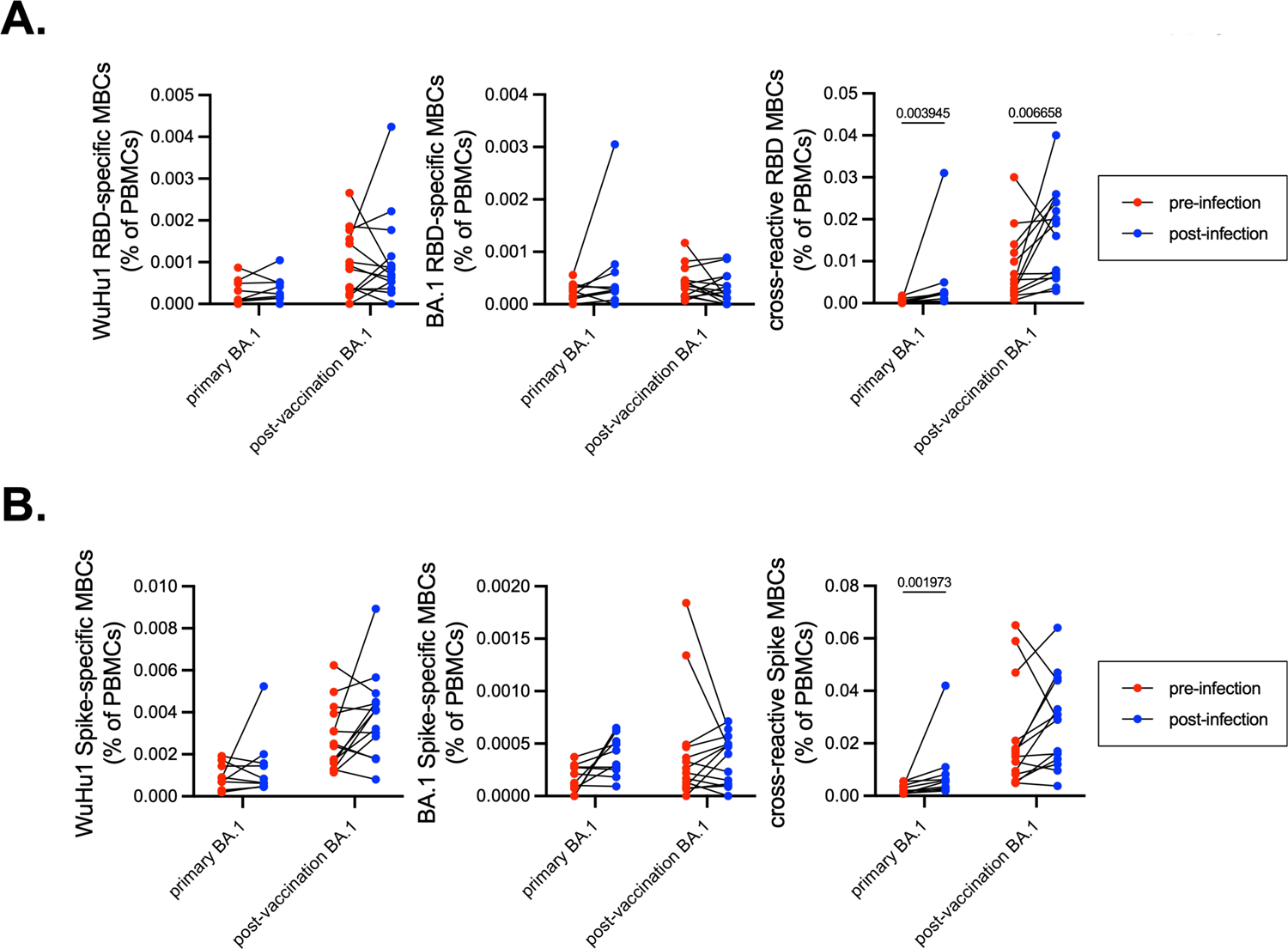
Frequency of WuHu1- and BA.1-specific memory B cells before and after BA.1 infection. **(A)** Frequencies of isotype-switched memory B cells with Wuhu1 RBD+, BA.1 RBD+ and cross-reactive RBD+ specificities in both unvaccinated and vaccinated individuals before and after BA.1 infection. Lines connect the same individual from pre-infection frequency to post-infection frequency. In primary infections, pre-infection blood draws were taken on average 75.6 days before infection and post-infection blood draws occurred on 37.8 days after infection. In post-vaccination infections, pre-infection blood draws were taken on average 87.6 days before infection and post-infection draws were taken an average of 38.3 days after infection. Individuals that received a vaccine after the pre-infection draw were excluded from analysis. P values were calculated using Wilcoxon matched-pairs signed rank test on each row and post hoc testing for multiple comparisons between draws was performed using two-stage linear step-up procedure of Benjamini, Krieger and Yekutieli. P values greater than 0.05 are not depicted. **(B)** Frequencies of isotype-switched memory B cells with Wuhu1 Spike+, BA.1 Spike+ and cross-reactive Spike+ specificities in both unvaccinated and vaccinated individuals before and after BA.1 infection. Lines connect the same individual from pre-infection frequency to post-infection frequency. P values were calculated using Wilcoxon matched-pairs signed rank test on each row and post hoc testing for multiple comparisons between draws was performed using two-stage linear step-up procedure of Benjamini, Krieger and Yekutieli. P values greater than 0.05 are not depicted.

To infer potential mechanisms of antigenic imprinting from these samples, we first correlated pre-infection cross-reactive Spike-specific memory B cells and post-infection BA.1 Spike-specific memory B cells. A negative correlation could indicate detrimental imprinting, whereby pre-existing memory B cells outcompete naïve B cells and inhibit the generation of variant-specific responses. Instead, we observed a slight positive but non-statistically significant correlation between pre-infection cross-reactive Spike-specific memory B cells and post-infection BA.1 Spike-specific memory B cells (**Figure 7A**). Similarly, we observed a non-significant positive correlation between post-infection cross-reactive Spike-specific memory B cells and post-infection BA.1 Spike-specific memory B cells (**Figure 7B**).

**Figure 7.**
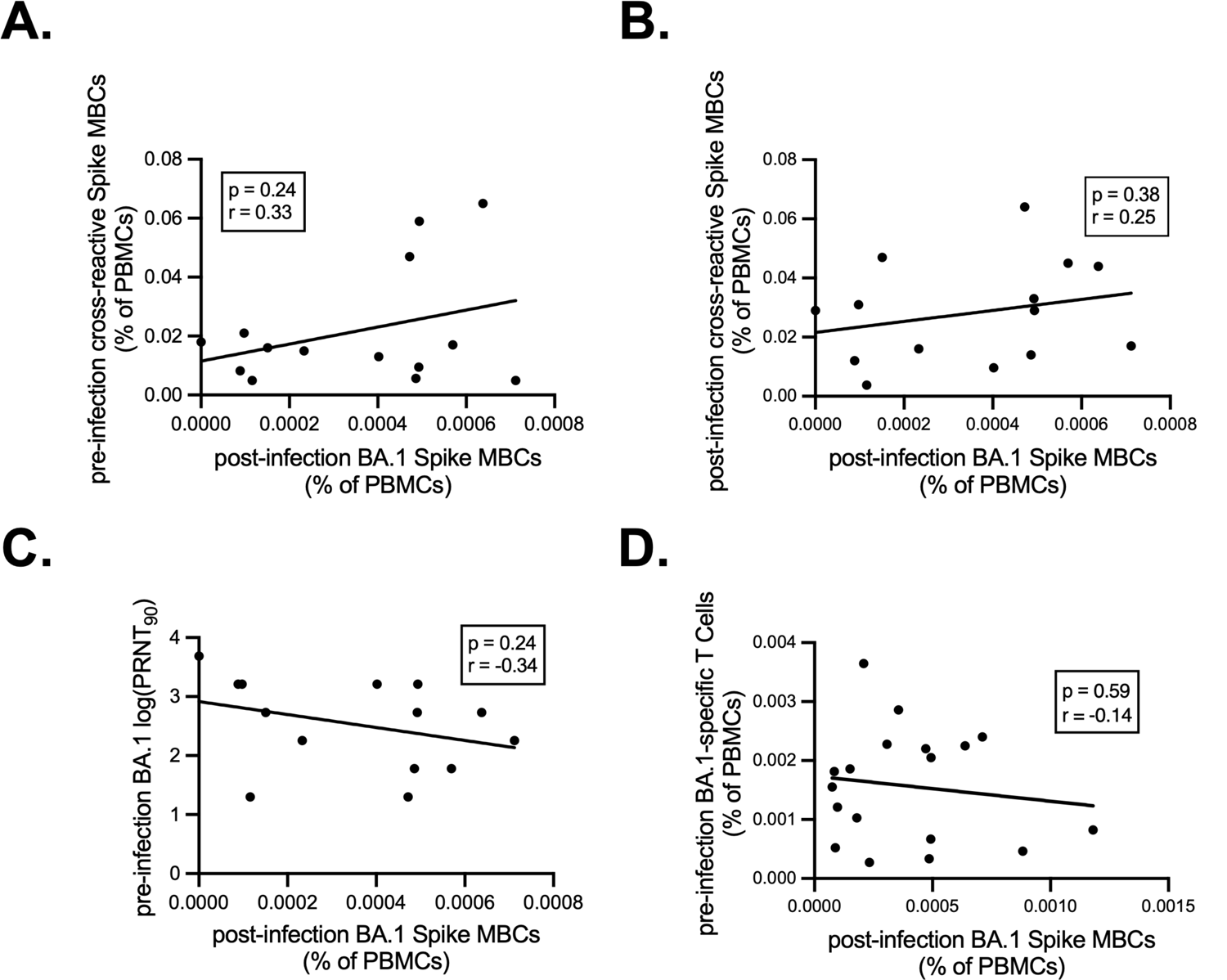
Correlations of pre-infection and post-infection BA.1-specific antibody, T and B cell responses. **(A)** Correlation of pre-infection cross-reactive Spike MBCs (calculated as in Figure 5C) plotted against the frequency of post-infection BA.1 Spike MBCs (calculated as in Figure 5C) in individuals that experienced a post-vaccination BA.1 infection. Pearson correlation analysis was performed. Pre-infection blood draws were taken on average 87.6 days before infection and post-infection draws were taken an average of 38.3 days after infection. Individuals that received a vaccine after the pre-infection draw were excluded from analysis. **(B)** Correlation of post-infection cross-reactive Spike MBCs (calculated as in Figure 6B) plotted against the frequency of post-infection BA.1 Spike MBCs (calculated as in Figure 5C) in individuals that experienced a post-vaccination BA.1 infection. Pearson correlation analysis was performed. **(C)** Correlation of pre-infection BA.1 neutralizing antibody titer (calculated as in Figure 4a) plotted against post infection BA.1 Spike MBCs (calculated as in Figure 5c) in individuals that experienced a post-vaccination BA.1 infection. Pearson correlation analysis was performed. **(D)** Correlation of pre-infection BA.1 Spike-specific T cells as measured by IFNγ ELISPOTs plotted against post-infection BA.1 Spike MBCs in individuals that experienced a post-vaccination BA.1 infection. Pearson correlation analysis was performed.

Given that these data do not support a mechanism of competitive inhibition of naïve B cells by cross-reactive memory B cells, we explored other mechanisms by which *de novo* responses to drifted epitopes are indirectly suppressed, such as accelerated viral clearance by neutralizing antibodies and/or T cells. We found a negative, but non-statistically significant correlation of *de novo* responses with pre-infection BA.1 neutralizing antibody titers (**Figure 7C**). Similarly, we observed a small and non-significant negative correlation with pre-infection BA.1 Spike-specific T cell numbers and post-infection BA.1 Spike memory B cells (**Figure 7D**). The small sample sizes and variable times of blood sampling prior to infection preclude us from making definitive conclusions about mechanisms driving antigenic imprinting. Nonetheless, the data suggest that neutralizing antibody and/or memory T cell-mediated viral clearance may indirectly underlie suppression of responses to drifted epitopes. This overall impact is quite small relative to the marked changes in antibody immunodominance observed in even primary BA.1 variant infections, irrespective of prior immunity.

## Discussion

Antigenic imprinting is neither inherently beneficial nor detrimental; rather the impact of prior immunity is context-dependent^45^. For example, pre-existing serum antibodies can improve and focus *de novo* responses upon reinfection to only mutated novel epitopes through epitope masking^104–106^. Similarly, *de novo* responses to drifted epitopes can be improved by pre-existing CD4+ memory T cells in what is classically known as the hapten-carrier effect^107^. Alternatively, high affinity memory B cells can competitively inhibit naïve B cells by consuming limited amounts of antigen and T cell help, leading to a suppression of *de novo* antibody responses^108^. If these memory B cells target non-protective epitopes, this could in theory leave one worse off than if there were no prior immunity whatsoever^44,109^. Finally, pre-existing immunity could indirectly suppress new antibody responses to drifted epitopes simply by clearing away virus and antigen before naïve B cells can robustly participate.

Though neutralizing antibody titers were robust following post-vaccination infections, our results demonstrated a small negative impact of prior immunity on *de novo* responses to drifted epitopes. Yet we found no evidence to support a mechanism of competitive inhibition by cross-reactive memory B cells. Though not definitive, our data instead hint at a role for pre-infection neutralizing antibodies and memory T cells, suggesting that antigen clearance is the main mechanism by which *de novo* B cell responses are indirectly suppressed by prior immunity. Indeed, pre-existing neutralizing antibodies likely accelerate viral clearance^110,111^, and viral and vaccine antigens can potentially also be cleared by T cells or non-neutralizing antibodies via Fc effector functions^112–114^. Animal studies offer an attractive way to further test mechanisms of antigenic imprinting on heterologous vaccine and viral infection responses. For example, genetic tracking studies were used to show robust *de novo* responses to Omicron boosters in mice previously vaccinated against the ancestral strain. Yet this required two booster doses, and a small negative impact of prior immunity was observed in inverse proportion to the antigenic distance between the two immunizations^101^. Similar results have been reported in other mouse studies^115^. These systems can thus potentially be used to manipulate specific immune parameters and measure their contributions to antigenic imprinting in ways that are not possible in human studies, especially since few immunologically naïve adults remain to serve as controls.

Immunodominance hierarchies can also determine which epitopes are available to be targeted by antibodies, irrespective of prior immunity. Prior studies, confirmed in our experiments, showed that a large portion of COVID-19 vaccine-induced antibodies are aimed at the L452 class 3 epitope^116^. Yet in the post-vaccination Delta cohort, we observed few antibodies directed at the epitope containing the L452R mutation. Under the assumption that one immunodominant epitope was being mutated to another, one might have concluded that the absence of R452-specific antibodies could be explained by antigenic imprinting. Yet by including a primary infection cohort, we observed that the Delta variant intrinsically did not elicit detectable antibody responses against the R452 epitope, even with no prior SARS-CoV-2 exposures, consistent with an independent study^52^. We can instead conclude that Delta shifts antibody immunodominance hierarchies to instead focus more on epitopes located in the NTD. These types of shifts in immunodominance preempt any considerations of the impact of antigenic imprinting. The basis and mechanisms of these shifts for SARS-CoV-2 clearly needs more investigation to determine whether and how best to overcome them.

This study spanned a period from the Delta wave through the more antigenically distinct BA.1 Omicron wave. A central expectation of antigenic imprinting is that the extent to which prior immunity interferes with *de novo* responses should decrease as antigenic distance increases^101^. We used the Delta and BA.1 variants to test this expectation in SARS-CoV-2 and to understand the impacts of antigenic distance on antigenic imprinting. Despite our prediction, we observe even less of a variant specific response in post-vaccination BA.1 infections compared to post-vaccination Delta infections. Much of this can be explained by shifts in immunodominance in which even primary BA.1 infections elicited few memory B cell responses to drifted epitopes. Yet longitudinal sampling during BA.1 infections has also shown that viral titers do not reach the peak levels observed in Delta infections^117^, suggesting that immune responses to drifted epitopes occur in proportion to need and antigen availability.

### Disclosures

The findings and conclusions in this report are those of the authors and do not necessarily represent the official position of the Centers for Disease Control and Prevention.

## Data Availability

All data produced in the present study are available upon reasonable request to the authors

https://www.ncbi.nlm.nih.gov/geo/query/acc.cgi?acc=GSE242775

## Acknowledgements

This work was supported by NIH grants R01AI099108 and R01AI129945 (D.B.) and a research grant from the Arizona Board of Regents (M.W. and D.B). This project has been funded in part with Federal funds from the National Institute of Allergy and Infectious Diseases, National Institutes of Health, Department of Health and Human Services, under Contract No. 75N93021C00015 (M.W.) The HEROES-RECOVER cohort is supported by the National Center for Immunization and Respiratory Diseases and the Centers for Disease Control and Prevention (contracts 75D30120R68013 to Marshfield Clinic Research Institute, 75D30120C08379 to the University of Arizona, and 75D30120C08150 to Abt Associates).

## Methods

### Participant selection

All human studies conducted at The University of Arizona were approved by the Institutional Review Board for the Human Subjects Protection Program^†^. Individuals who had participated in the voluntary on-campus saline gargle testing program and had either never tested positive or had tested positive during the Delta or BA.1 waves were contacted by email by the program administrators (not the authors on this study) about willingness to participate in this research study. Participants were provided a link to an eligibility questionnaire and, once eligibility (no immunosuppressive therapy in the last 5 years and HIV negative) was confirmed, additional demographic questions and a link to schedule an appointment for blood draws. Written consent was obtained through an electronic form. All blood draws were performed at the Clinical and Translational Sciences Center at The University of Arizona. Additional primary and post-vaccination BA.1 infection samples were acquired from the CDC HEROES-RECOVERS^††^ cohort^85^. This study was reviewed by CDC and approved by the institutional review boards at participating sites or under a reliance agreement with Abt Associates institutional review board and was conducted consistent with applicable federal law and CDC policy under 45 C.F.R. part 46, 21 C.F.R. part 56, 42 U.S.C. Sect. 241(d), 5 U.S.C. Sect. 552a, 44 U.S.C. Sect. 3501 et seq. Methods for the HEROES-RECOVER Cohorts have been published previously^85,86^. In summary, cohorts consisted of health care personnel, first responders, and other essential and frontline workers in eight U.S. locations across six states. Participants collected weekly nasal swabs which were tested for SARS-CoV-2 viral material by RT-qPCR and additional swabs were collected and screened upon the onset of any COVID-19–like illness symptoms. In addition, blood draws were collected at enrollment, then approximately every 3 months and after immune modifying events such as vaccination or infection. Vaccination was documented by self-report and verified by vaccine cards or electronic medical records or state immunization registries. HEROES-RECOVER participants were selected based on testing positive for SARS-CoV-2 during Delta or BA.1 waves and having completed a blood draw after infection.

### Saline Gargle PCR testing for SARS-CoV-2

As part of Test All, Test Smart, the University of Arizona’s voluntary campus-wide testing program, University staff, faculty and students had access to SARS-CoV-2 rRT-PCR tests from August 2020 – July 2023. At testing and collection sites throughout campus, individuals were given 5 mL of 0.9 % sterile saline (AddiPak 5 mL sterile saline single use tubes, Teleflex, LLC) and guided to complete three rounds of a 5-second swish followed by 10 seconds of gargling (adapted from Goldfarb et al.^118^). Samples were deposited into collection tubes and then screened for SARS-CoV-2 by rRT-PCR.

### PBMC and plasma preparation

Twenty milliliters of blood was collected by venipuncture in heparinized Vacutainer tubes (BD). For PBMCs, 15ml of Ficoll-Paque PLUS (Thermo Fisher Scientific) was added to 50-ml Leucosep tubes (Greiner) and spun for 1min at 1,000g to transfer the density gradient below the filter. Twenty milliliters of blood from the heparinized tubes was then poured into the top of the Leucosep tube and spun at 1,000g for 10min at room temperature with the brake off. The top plasma layer was carefully collected and frozen at −20 °C, and the remaining supernatant containing PBMCs above the filter was poured into a new 50-ml conical tube containing 10mL of PBS and spun at 250g for 10min. Cell pellets were resuspended in RPMI media containing 10% FCS and counted on a Vi-Cell XR (Beckman Coulter). Cells were diluted to a concentration of 2 × 10^!^ cells per mL in RPMI media containing 10% FCS. An equal volume of 80% FCS + 20% dimethyl sulfoxide was added dropwise and inverted once to mix. Suspensions were distributed at 1ml per cryovial and frozen overnight at −80 °C in Mr. Frosty freezing chambers (Nalgene). Vials were then transferred to storage in liquid nitrogen.

### ELISA and quantification of antibody titers

Serological assays were performed as previously described^88^. WuHu1 RBD (cat. no. SPD-C52H3), WuHu1 S1subdomain of the SARS-CoV-2 S glycoprotein (cat. no. S1N-C52H3), WuHu1 Spike (cat. no. SPN-C52H9), Delta RBD (cat. no. SPD-C52Hh), Delta S1 subdomain (cat. no. S1N-C52Hu), Omicron (BA.1) RBD (cat. no. SPD-C522e), Omicron (BA.1) Spike (cat. no. SPN-C52Hz) and Nucleocapsid (cat. no. NUN-C5227) were purchased from Acro Biosystems. Chimeric proteins (Delta RBD-L452 and Delta NTD-WuHu1 S1) were custom synthesized by GenScript. To obtain titers and single-dilution OD450 values, antigens were immobilized on high-adsorbency 384-well plates at 5 ng mL^−1^. Plates were blocked with 1% non-fat dehydrated milk extract (Santa Cruz Biotechnology, sc-2325) in sterile PBS (Thermo Fisher Scientific HyClone PBS, SH2035) for 1 h, washed with PBS containing 0.05% Tween-20 and overlaid for 60 min with either a single 1:60 dilution or five serial 1:3 dilutions beginning at a 1:60 dilution of serum. Plates were then washed and incubated for 1 h in 1% PBS and milk containing anti-human Pan-Ig HRP-conjugated antibody (Jackson ImmunoResearch, 109-035-064) at a concentration of 1:2,000 for 1 h. Plates were washed with PBS-Tween solution followed by PBS wash. To develop, plates were incubated in tetramethylbenzidine (Fisher Scientific) before quenching with 2 N H_2_SO_4_. Plates were read for 450-nm absorbance on CLARIOstar Plus from BMG Labtech. All samples were also read at 630 nm to detect any incomplete quenching. Any samples above background 630-nm values were re-run. Area under the curve (AUC) values were calculated in GraphPad Prism (v9).

### Virus neutralization assays

All live virus assays were performed at Biosafety Level 3 and were approved by the University of Arizona Institutional Biosafety Committee. SARS-CoV-2, isolate USA-WA1/2020, was deposited by Dr Natalie J. Thornburg at the Centers for Disease Control and Prevention and obtained from the World Reference Center for Emerging Viruses and Arboviruses. Stocks of WA1/2020 SARS-CoV-2 were generated as a single passage from received stock vial on mycoplasma-negative Vero cells (ATCC CCL-81). B.1.617.2 (Delta) was received from WRCEVA, strain designation GNL-1205. B.1.1.529 (Omicron) originated from a nasopharyngeal swab collected at the University of Arizona. It was passaged once on Calu-3 cells and then once on Vero cells to generate a master stock. Viral PANGO-lineage, BA.1.1^87^, was confirmed by Illumina sequencing (EPI_ISL_17886211) of the master stock.

Supernatant and cell lysate were combined, subjected to a single freeze–thaw and then centrifuged at 1,800g for 10min to remove cell debris. For PRNTs for SARS-CoV-2, Vero cells (ATCC, CCL-81) were plated in 96-well tissue culture plates and grown overnight. Vero cells were confirmed by PCR to be free of mycoplasma using the Universal Mycoplasma Detection Kit (ATCC). Serial dilutions of serum samples were performed in duplicate and incubated with 100 plaque-forming units of SARS-CoV-2 for 1h at 37 °C. Plasma/serum dilutions plus virus were transferred to the cell plates and incubated for 2h at 37 °C in 5% CO2 and then overlaid with 1% methylcellulose. After 72h, plates were fixed with 10% neutral buffered formalin for 30min and stained with 1% crystal violet. Plaques were imaged using an ImmunoSpot Versa plate reader. The most dilute serum concentration that led to ten or fewer plaques was designated as the PRNT90 titer. Input PFU for each experiment was confirmed by plaque assay.

### Flow cytometry

One milliliter of pre-warmed FBS was added to a frozen cryovial of 10^7^ PBMCs, which was rapidly thawed in a 37℃ water bath. Samples were poured into 15 mL conical tubes containing 5 mL of pre-warmed RPMI with 5% FBS and 1% anti/anti. Tubes were spun at 250g for 5 min at room temperature.

### Delta

Supernatants were removed and cell pellets were resuspended in 200 µL of staining buffer containing 1 µL each of anti-CD38-APC (BioLegend, clone HIT2), anti-CD13-PE-Cy7 (BioLegend, clone WM15), anti-CD21-PE-Dazzle (BioLegend, clone Bu32), anti-CD19-APC-efluor-780 (Invitrogen, clone HIB19), anti-IgD-PerCP-Cy5.5 (Biolegend, clone IA6-2), anti-IgM-FITC (Biolegend, clone MHM-88), anti-CD27-BV510 (Biolegend, clone M-T271), anti-CD11c-Alexa700 (BioLegend, clone Bu15). Staining buffer also contained Delta-S1-PE and S1-BV421 tetramers.

### BA.1

Supernatants were removed and cell pellets were resuspended in 200 µL of staining buffer containing 1 µL each of anti-CD38-BV421 (BioLegend, clone HB-7), anti-CD13-PE-Dazzle 594 (BioLegend, clone WM15), anti-CD21-PerCP Cy 5.5 (BioLegend, clone Bu32), anti-CD19-APC-efluor-780 (Invitrogen, clone HIB19), anti-IgD-BV510 (Biolegend, clone 11-26c.2a), anti-IgM-FITC (Biolegend, clone MHM-88), anti-CD27-PE Cy 7 (Biolegend, clone M-T271), anti-CD11c-Alexa700 (BioLegend, clone Bu15). Cells were stained with live-dead marker, Zombie Yellow (BioLegend) according to manufacturer’s recommendations. Staining buffer also contained BA.1-Spike-PE and Spike-Alexa Fluor 647 tetramers.

Antibodies were validated by the manufacturer on human PBMCs. Tetramer reagents were assembled by mixing 100µg ml^-1^ of C-terminal AviTagged S1, Delta S1, WuHu1 Spike, or BA.1 Spike (ACROBiosystems) with 100µg ml^-1^ of streptavidin-PE(BioLegend), streptavidin-BV421 (BioLegend), or streptavidin-Alexa Fluor 647 (BioLegend), respectively, at a 6:1 molar ratio for S1 or 4:1 molar ratio for Spike, in which ⅕ of the final volume of streptavidin was added every 10 min. S1 and Spike tetramers were validated by staining Lenti-X 293T cells(Takara Bio) as a negative control or 293T-hACE2-expressing cells (BEI Resources, NR-52511) as a positive control. Lenti-X 293T cells were confirmed to be free of mycoplasma; 293T-hACE2 cells were maintained in media containing 1% pen/strep to minimize chances of contamination. PBMC samples were stained for at least 20 minutes, washed and filtered through 70-µm nylon mesh. Data were analyzed on either a BD LSR2 (tetramer validation only), a Fortessa cytometer (Delta), or BD Cytek Aurora (BA.1). Data were analyzed using FlowJo software.

### Flow cytometry and Fluorescence Activated Cell Sorting

One milliliter of pre-warmed FBS was added to a frozen cryovial of PBMCs and thawed by pipetting. Samples were added to 15 mL conical tubes containing 10 mL of pre-warmed RPMI with 20% FBS and 1% anti/anti. Tubes were spun at 1200 RPM for 5 minutes at room temperature. Supernatants were removed and cell pellets were resuspended in 200 µL of staining buffer contained 1 µL each of anti-CD19-BV421(Biolegend, clone HIB19), anti-CD27-FITC(Biolegend, clone O323), anti-CD13-PE-Cy-7(Biolegend, clone WM15), anti-IgD-APC-Cy-7(Biolegend, clone IA6-2). Staining buffer also contained either 5 or 2 LIBRA-Seq tetramers: S1-PE(Biolegend, TotalSeq-C0951_PE), Delta S1-PE(Biolegend, TotalSeq-C0952_PE), Delta NTD/S1-PE(Biolegend, TotalSeq-C0953_PE), Delta RBD/L452(Biolegend, TotalSeq-C0954_PE), and Delta RBD-PE(Biolegend, TotalSeq-C0955_PE). Biotinylated tetramer reagents were assembled by mixing 100µg ml^-1^ of C-terminal AviTagged S1 (ACROBiosystems), Delta S1 (ACROBiosystems), Delta NTD/S1 (GenScript), Delta RBD/L452 (Genscript), or Delta RBD (ACROBiosystems) with 100µg ml^-1^ of streptavidin-TotalSeq-C-PE (BioLegend) at a 6:1 molar ratio in which ⅕ of the final volume of streptavadin was added every 10 min. Tetramers were validated by staining Lenti-X 293T cells(Takara Bio) as a negative control or 293T-hACE2-expressing cells (BEI Resources, NR-52511) as a positive control. Lenti-X 293T cells were confirmed to be free of mycoplasma; 293T-hACE2 cells were maintained in media containing 1% pen/strep to minimize chances of contamination. Additionally, TotalSeq-C anti-human Hashtag antibodies (Biolegend, TotalSeq™-C0251-10) were added to individual samples and pooled after staining and washing. PBMCs were stained in the dark for 30 minutes at 4°C, washed, pooled and filtered through a 35 µm strainer (Fisher Scientific). SARS-CoV-2 specific memory B cells (CD19+IgD-IgM-CD27+) as well as non-antigen specific memory B cells were sorted using a FACSAria II.

### Single-cell RNA sequencing and analysis

Cells were prepared and processed according to the 10X Genomics Single Cell 5’ Dual Index protocol with Feature Barcoding Technology for Cell Surface Protein and Immune Receptor Mapping kit (10X Genomics). Reads were processed and aligned using the 10X CellRanger multi pipeline to GRCh38 gex and vdj reference genomes (10X Genomics). Each sample feature barcode matrix was loaded into R and analyzed utilizing the Seurat package for gene expression, vdj and antibody capture analysis^119^. Cell processing was conducted as previously described^100^. Data are available at NCBI GEO accession number GSE242775.

### ELISpot Assay

Cryopreserved PBMC (5 × 10^6^/sample) were thawed in prewarmed RPMI-1640 media supplemented with L-glutamine + 10% FCS and 300ug DNAse. Thawed PBMCs were rested overnight at 37 °C in X-VIVO™-15 Medium (Lonza) supplemented with 5% human-AB serum. Cells were stimulated with ∼1 nmol of peptide pool corresponding to spike of Omicron (B.1.1.529) variant (16-mer peptide pools, overlapping by 10 amino acids (21st century Biochemicals Inc.) on pre-coated human IFN-γ ELISpot plates (Mabtech, Inc.) and developed after 18 hours according to manufacturer instructions. Spots were imaged and counted using Iris FLUOROspot reader (Mabtech).

### Statistical methods

All analyses are listed in the figure legends and were performed in GraphPad Prism 9 and/or the R programming language (v4.0.5).

### Footnotes

^†^ See 45 C.F.R. part 46; 21 C.F.R. part 56

^††^ This study was reviewed by CDC and approved by the institutional review boards at participating sites or under a reliance agreement with Abt Associates institutional review board and was conducted consistent with applicable federal law and CDC policy under 45 C.F.R. part 46, 21 C.F.R. part 56, 42 U.S.C. Sect. 241(d), 5 U.S.C. Sect. 552a, 44 U.S.C. Sect. 3501 et seq.

---

**Figure S1.**
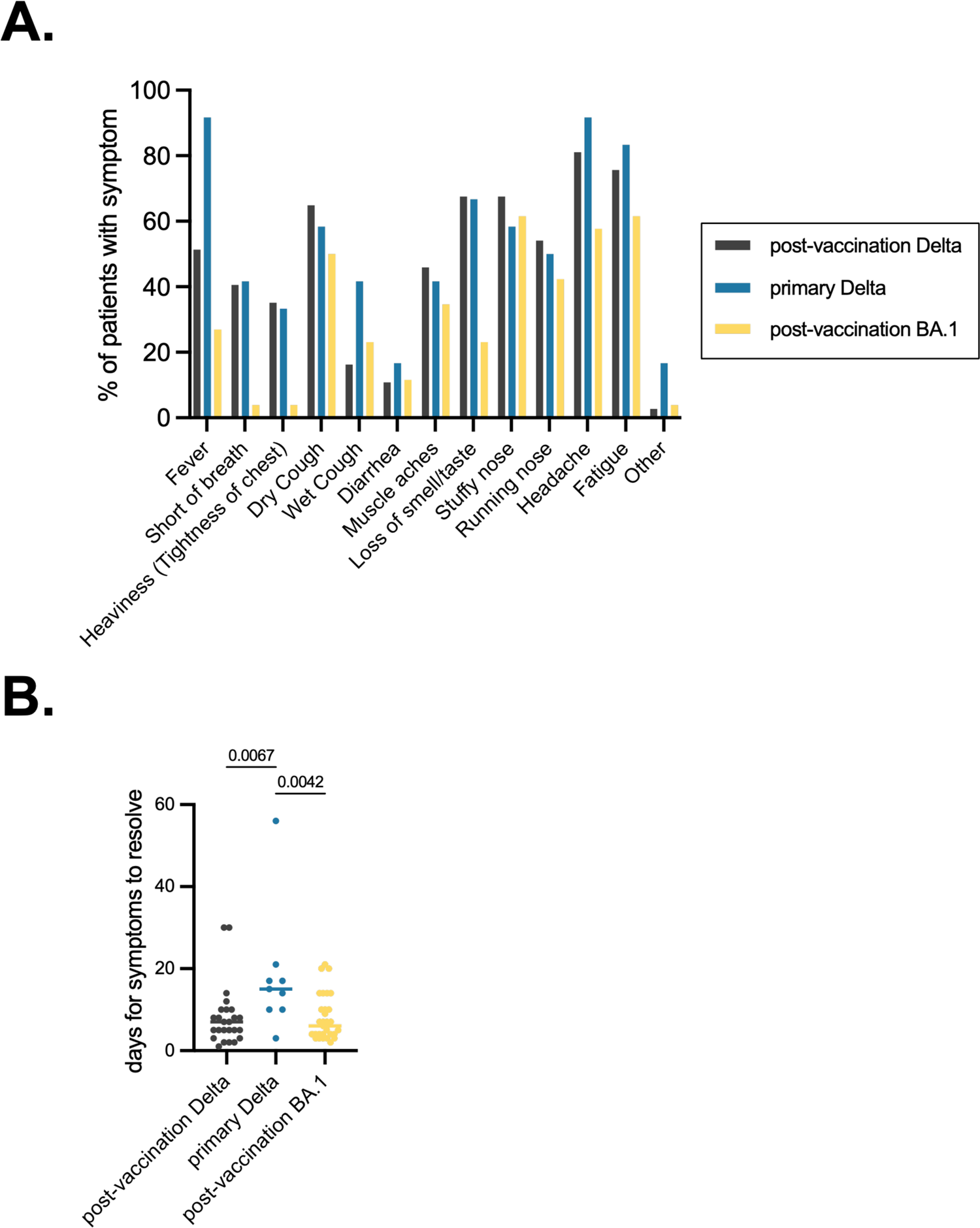
Test All, Test Smart (TATS) symptom report. **(A)** Percentage of individuals from each TATS cohort that reported experiencing various respiratory/cold symptoms in study entry survey. **(B)** Reported days until symptoms resolved for each TATS cohort. Two-sided P values from t-test statistics were calculated for pairwise differences using one-way ANOVA. Post hoc testing for multiple comparisons between draws was performed using Tukey’s multiple comparisons test. P values greater than 0.05 are not depicted.

**Figure S2.**
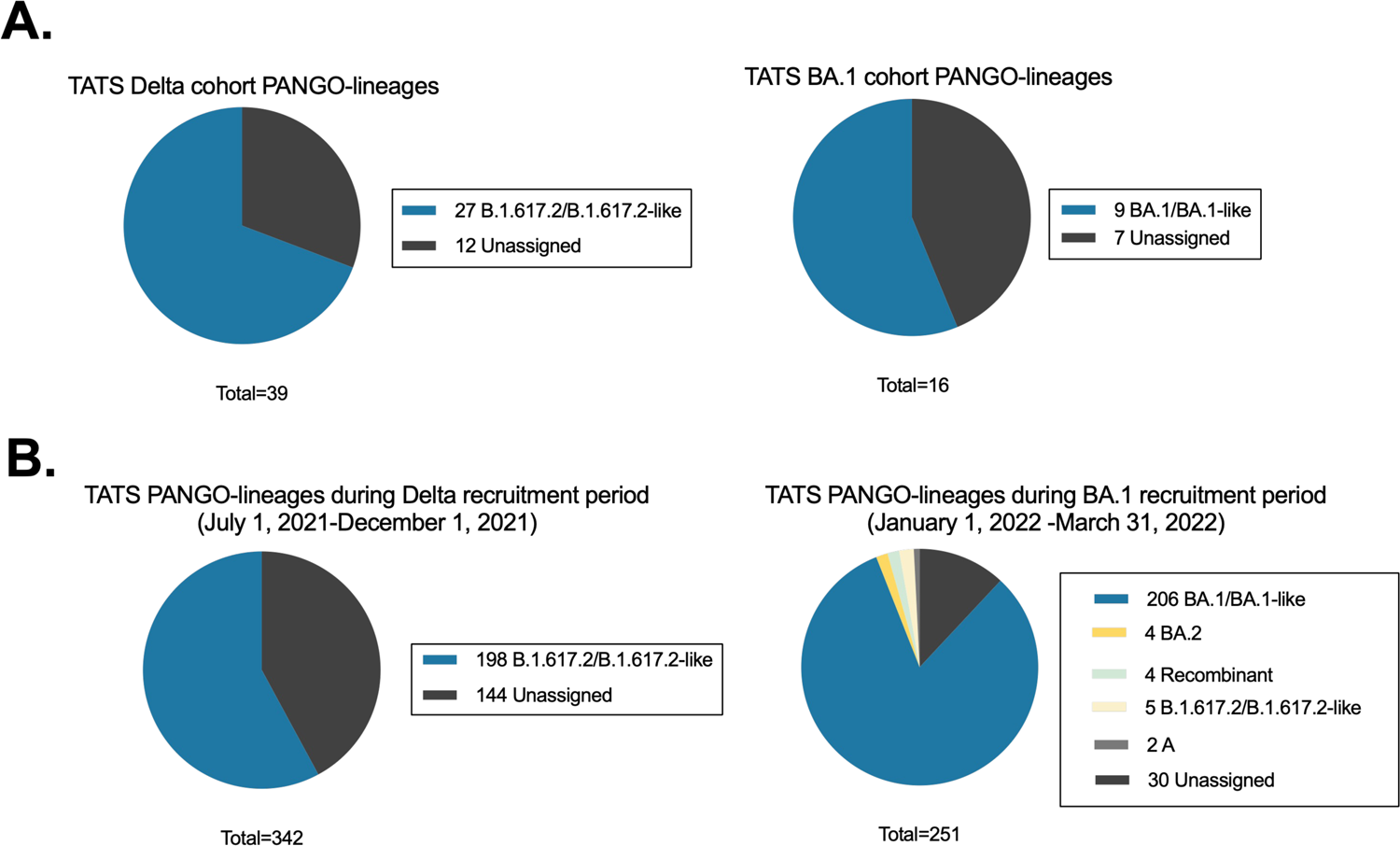
PANGO-lineage assignments from TATS PCR positive individuals. **(A)** Delta or BA.1 PANGO-lineage assignments after SARS-CoV-2 viral amplicon sequencing (Integrated DNA Technologies). Unassigned sequences could not be assigned to a PANGO-lineage due to insufficient viral RNA recovery and low sequence coverage. **(B)** PANGO-lineage assignments of all TATS samples submitted during the period of Delta cohort recruitment, July 1, 2021-December 1, 2021 (left panel) or during the period of BA.1 cohort recruitment, January 1, 2022-March 31, 2022 (right panel). Unassigned sequences could not be assigned a lineage due to insufficient viral RNA recovery and low sequence coverage.

**Figure S3.**
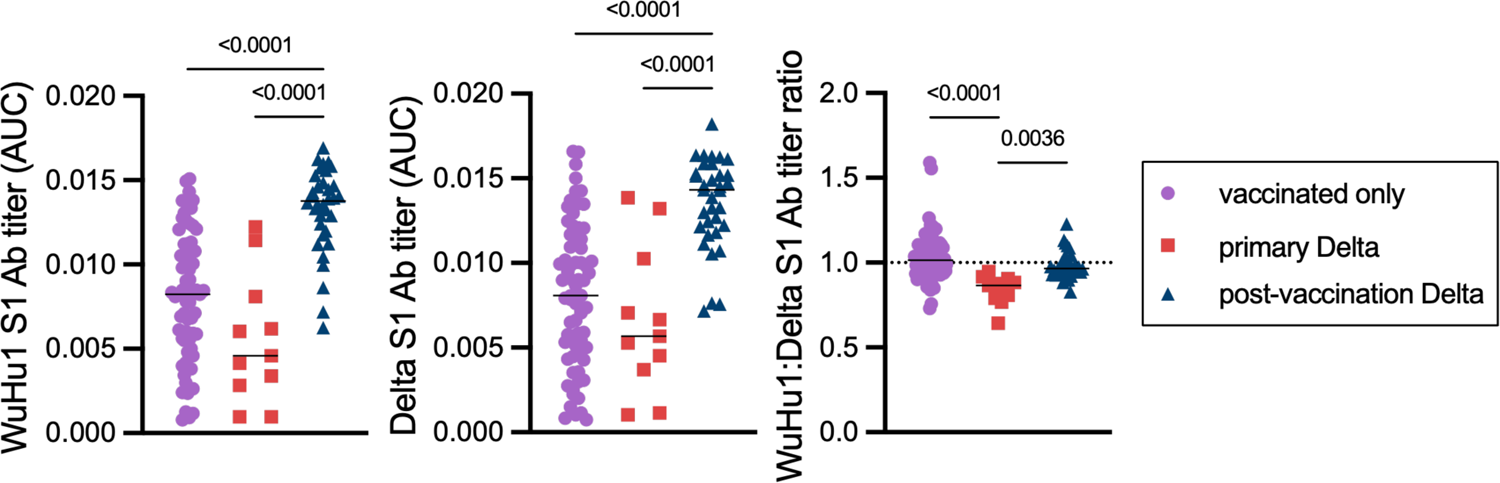
Primary and recall antibody responses to Wuhan and Delta strains of SARS-COV-2. Quantitative titers of WuHu1- and Delta S1-specific antibodies. Serum was initially diluted 1:60, serially diluted 1:3, assessed by ELISA for binding to the listed antigens, and area under the curve (AUC) values were calculated. Each symbol represents an individual. WuHu1 AUC values were divided by their Delta AUC titer in the same individual to calculate a WuHu1:Delta S1 ratio in the rightmost panel. Two-sided P values from t-test statistics were calculated for pairwise differences using one-way ANOVA. Post hoc testing for multiple comparisons between draws was performed using Tukey’s multiple comparisons test. P values greater than 0.05 are not depicted.

**Figure S4.**
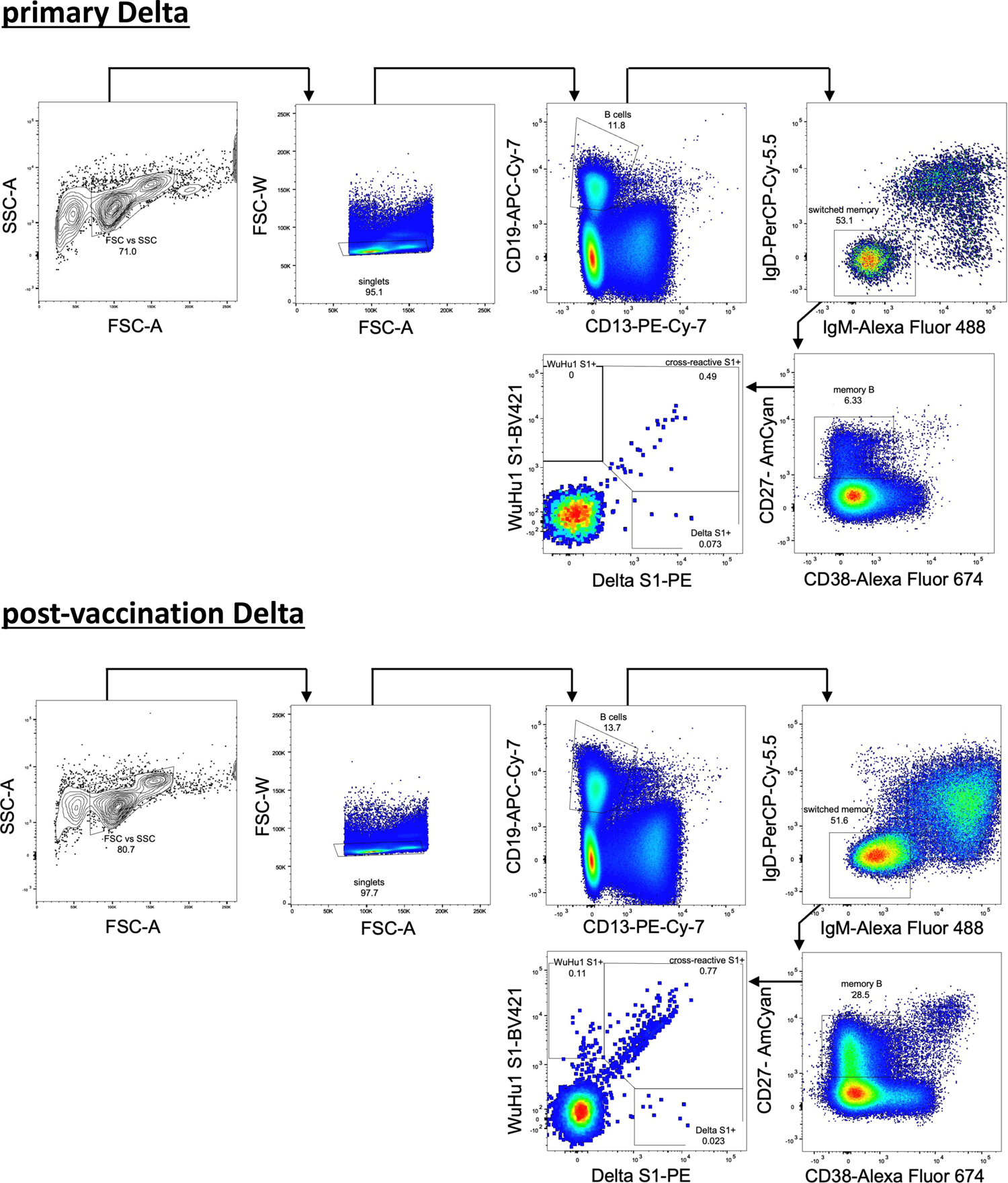
Flow cytometric gating strategy with Delta S1 and WuHu1 S1 tetramers. Examples of a sample from a primary Delta infection (top) and post-vaccination Delta infection (bottom) are shown.

**Figures S5.**
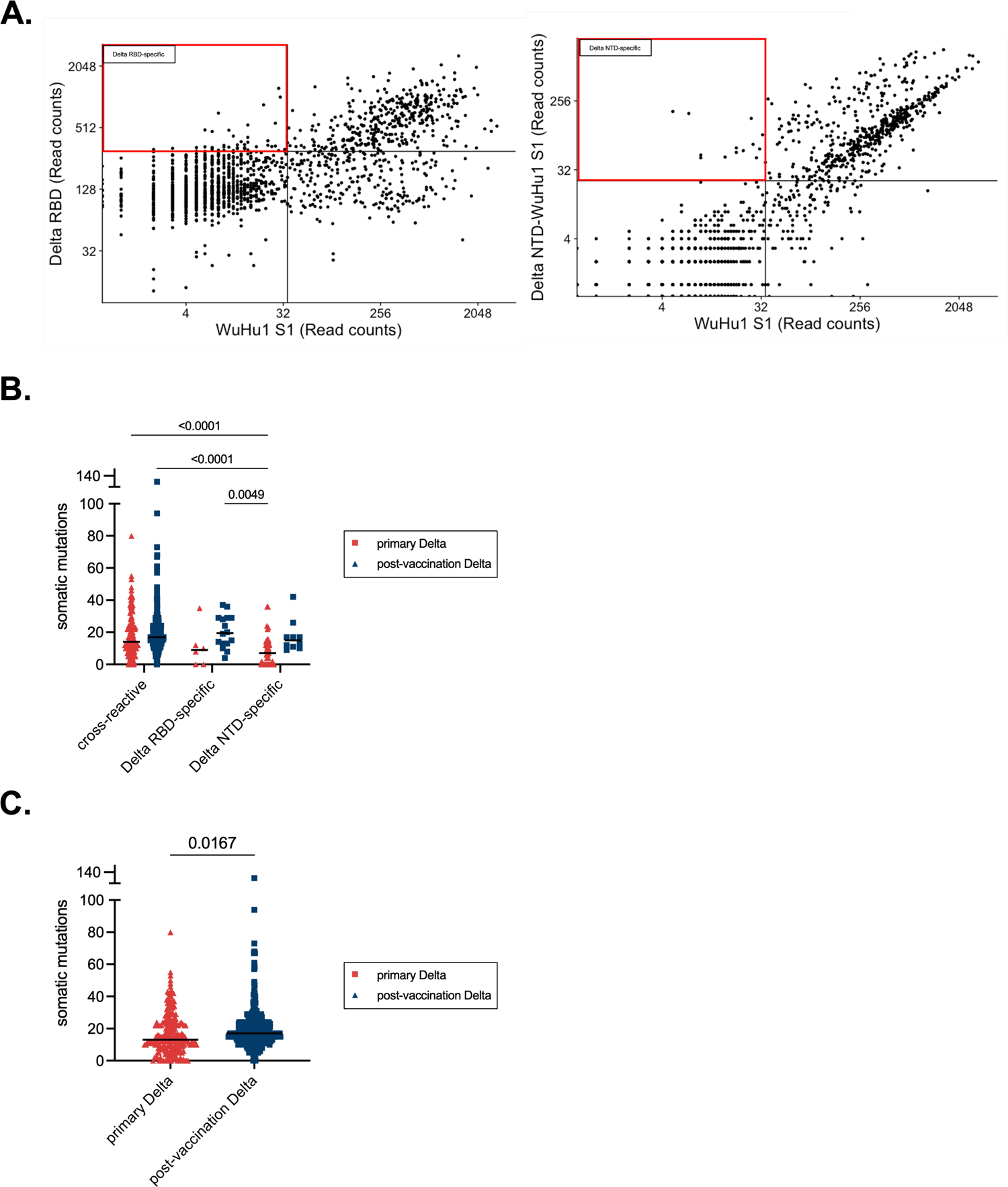
LIBRA-seq analysis in primary and post-vaccination Delta infections and quantification of somatic mutations. **(A)** A chimeric protein (Delta NTD-WuHu1 S1) was generated in which Delta NTD mutated epitopes (T19R, G142D, E156-, F157-, R158G) were incorporated into the otherwise WuHu1 S1 backbone. Quantification of Delta RBD-specific **(left)** and Delta NTD-specific memory B cells **(right)** in individuals that experienced a post-vaccination Delta infection. Delta RBD-specific cells were classified by cells that had Delta RBD read counts of greater than 300 and WuHu1 S1 read counts of less than 35. Delta NTD-specific cells were classified by cells that had Delta NTD-WuHu1 S1 read counts of greater than 23 and WuHu1 S1 read counts of less than 35. Read count thresholds to determine positivity were set using samples in which cells lacking Spike-binding specificities were sorted and sequenced. Plots are concatenated from ten individuals. **(B)** Somatic mutations were calculated using the observedMutations command in the Shazam Immcantation package in R. Specificities of cells are determined using the same cutoffs described in **Figure S3A and 3D. (C)** Quantification of somatic mutations of all Spike specific cells subjected to scRNAseq from either ten primary or post-vaccination Delta infections.

**Figure S6.**
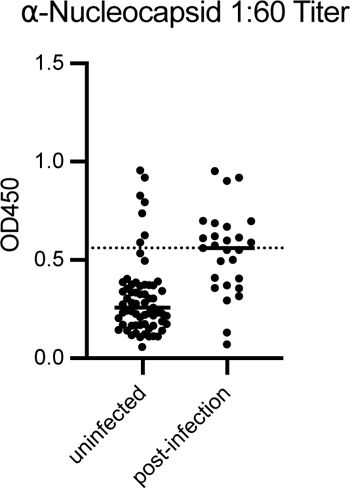
anti-Nucleocapsid titers in uninfected individuals. Individuals with ⍺-Nucleocapsid titers of greater than 0.6 at a1:60 serum dilution were considered previously infected and excluded from the study.

**Figure S7.**
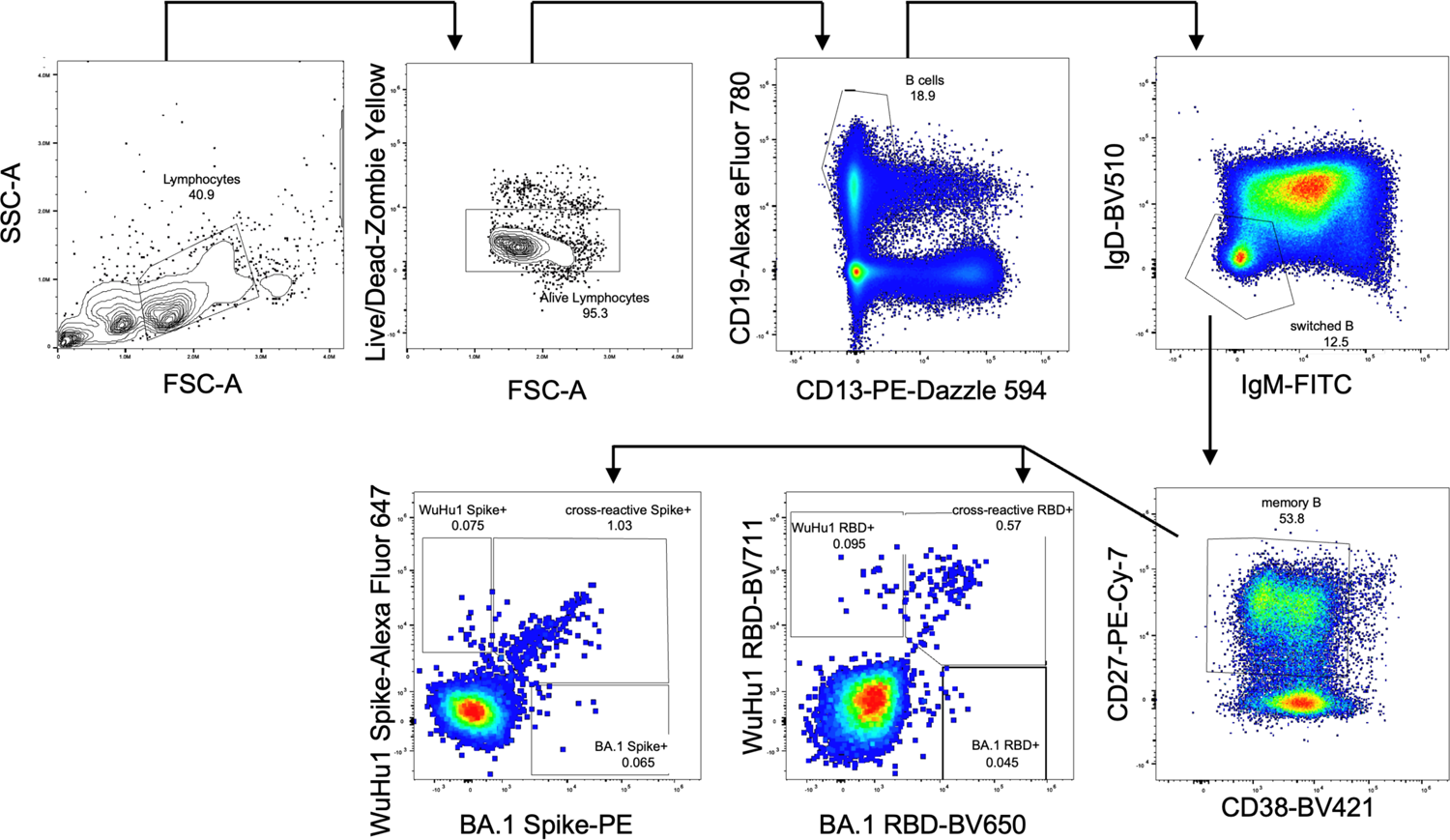
Flow cytometric gating strategy with BA.1 RBD, BA.1 Spike, WuHu1 RBD and WuHu1 Spike tetramers. An example of a sample from a post-vaccination BA.1 infection is shown.

**Figure S8.**
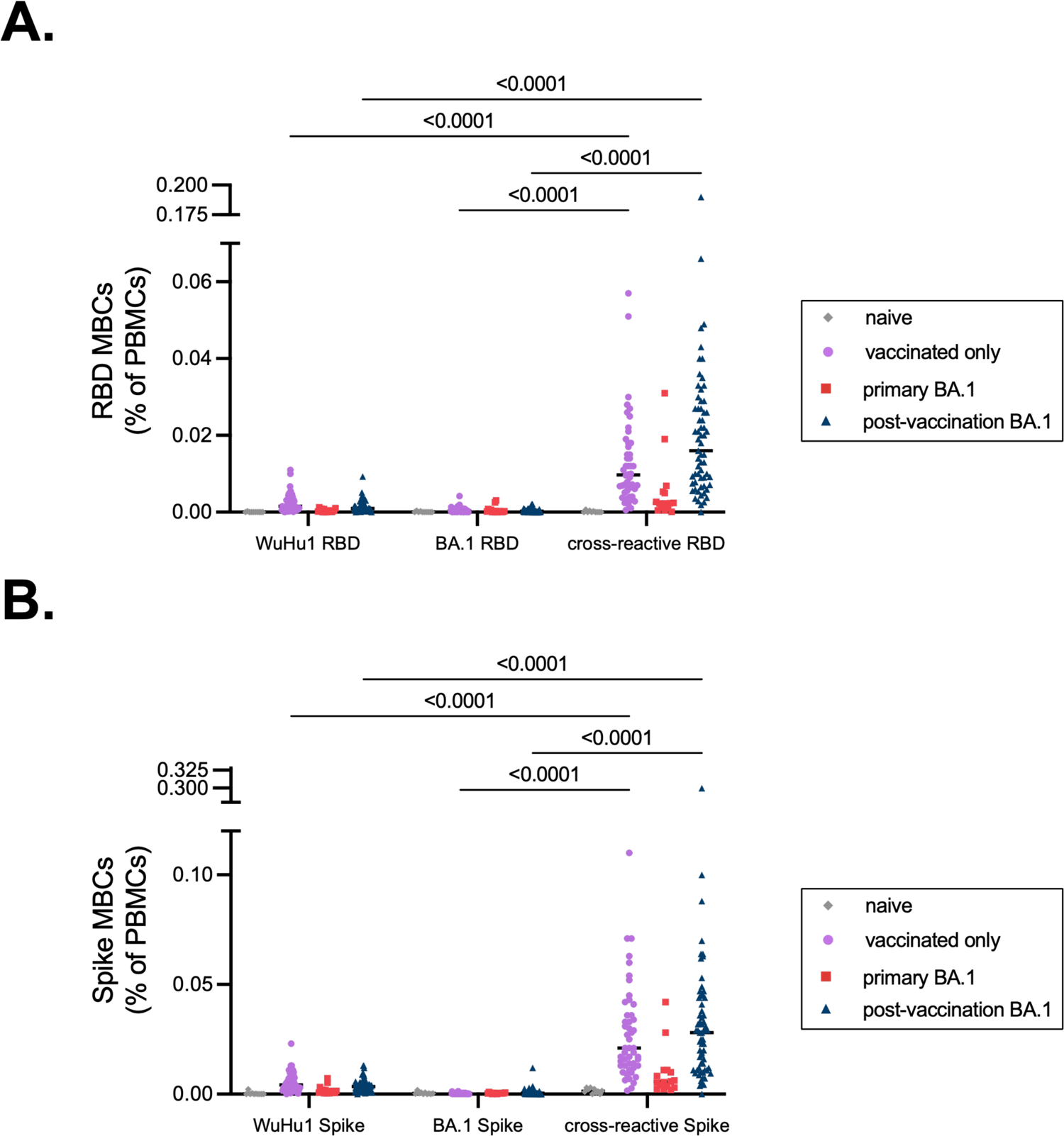
WuHu1 and BA.1 Memory B cell flow cytometric quantification. **(A)** Cells that bind both WuHu1 RBD and BA.1 RBD are annotated as cross-reactive RBD+, whereas cells that bind only WuHu1 RBD or BA.1 RBD are annotated as WuHu1 RBD+ or BA.1 RBD+, respectively. Quantification of isotype-switched memory B cells as a percentage of total PBMCs for Wuhu1 RBD+, BA.1 RBD+ and cross-reactive RBD+ specificities for each cohort of SARS-CoV-2 immune histories. Each symbol represents an individual. Two-sided P values from t-test statistics were calculated for pairwise differences using two-way ANOVA. Post hoc testing for multiple comparisons between draws was performed using Tukey’s multiple comparisons test. P values greater than 0.05 are not depicted. **(B)** Cells that bind both WuHu1 RBD and BA.1 Spike are annotated as cross-reactive Spike+, whereas cells that bind only WuHu1 Spike or BA.1 Spike are annotated as WuHu1 Spike+ or BA.1 Spike+, respectively. Quantification of isotype-switched memory B cells for Wuhu1 Spike+, BA.1 Spike+ and cross-reactive Spike+ specificities for each cohort of SARS-CoV-2 immune histories. Each symbol represents an individual. Two-sided P values from t-test statistics were calculated for pairwise differences using two-way ANOVA. Post hoc testing for multiple comparisons between draws was performed using Tukey’s multiple comparisons test. P values greater than 0.05 are not depicted.

